# Genome-wide association and gene-virus interaction study of liver disease in hepatitis C virus-infected patients

**DOI:** 10.1101/2025.10.12.25337816

**Authors:** Jocelyn Quistrebert, Haiting Chai, Yan Chen, Narayan Ramamurthy, Hamish Innes, Jennifer Benselin, Zhiqing Wang, Qijing Shen, Emanuele Marchi, Vincent Pedergnana, Paul Klenerman, Graham Cooke, Eleanor Barnes, William L. Irving, John McLauchlan, M. Azim Ansari

## Abstract

**Background and Aims:** Chronic hepatitis C (CHC) can progress to cirrhosis and hepatocellular carcinoma (HCC). This study aimed to identify genetic determinants and host-viral interactions that drive this progression to inform risk stratification and personalised treatment strategies.

**Methods:** We performed a genome-wide association study (GWAS) of cirrhosis (2,829 cases and 1,515 CHC controls), followed by a GWAS of HCC (706 cases and 2,152 cirrhosis controls). We performed cis-eQTL mapping and deconvolution in liver tissue of HCV-infected (136 CHC and 54 cirrhosis) patients to investigate gene expression regulation and cellular heterogeneity. Additionally, ten polygenic risk scores (PRS) for non-viral liver diseases were tested in 3,406 infected individuals.

**Results:** We identified the missense risk variant rs738409 in *PNPLA3* and a protective variant (rs4386418) in *XKR3* in genotype 1-infected patients that were significantly associated with cirrhosis but not HCC progression. HLA fine-mapping identified two amino acids in *HLA-DQB1*03:01* and *HLA-DRB1*13:01* associated with cirrhosis risk. No genome-wide significant association was observed for HCC, and loci previously linked to non-viral HCC did not replicate. The eQTL analysis revealed 2,060 genes under cis-regulatory control and 129 whose effects were modified by cirrhosis. An intronic eQTL lowered *PNPLA3* expression, but was not linked to cirrhosis risk. Deconvolution revealed expansion of plasma cells and macrophages and depletion of hepatocytes in CHC, with further immune-stromal remodelling in cirrhosis. All PRS showed a significant association with cirrhosis risk but not HCC progression.

**Conclusion:** Cirrhosis in CHC shares genetic architecture with non-viral liver diseases but also displays virus-specific risk variants. Cirrhosis risk involves genetic factors that differ from those underlying progression to HCC.

## Introduction

Chronic hepatitis C (CHC) infection is a leading cause of liver-related morbidity and mortality, with approximately 5–15% of patients progressing to cirrhosis over 20–40 years of infection^1,2,3^. Once cirrhosis is established, patients have a 1% annual risk of hepatocellular carcinoma (HCC)^4^. The progression from CHC to these severe liver conditions is extremely variable and influenced by a complex interplay of viral, environmental, and host genetic factors^1^. Genetic variations can influence the host’s immune response, viral clearance, and the extent of liver damage, thereby affecting the progression to various stages of liver disease^5^.

Although a sustained virological response (SVR) after direct-acting antiviral (DAA) treatment is associated with a reduction of liver morbidity and mortality, a residual risk of disease progression remains in some patients^6,7^. Identifying genetic variants that predispose individuals to liver disease could enhance our understanding of the disease’s biological mechanisms and aid in the development of targeted therapies independent of viral pathology.

Across non-viral liver disease, three common loci, PNPLA3 (p.I148M), TM6SF2 (p.E167K) and MBOAT7 (rs641738) are reproducibly associated with steatosis, fibrosis/cirrhosis and HCC risk, particularly in NAFLD and alcohol-related liver disease^8,9,10^. These established signals provide a benchmark to test whether HCV-related cirrhosis and HCC share pathways with non-viral aetiologies or reflect virus-specific mechanisms. Candidate gene studies and large-scale genome-wide association studies (GWASs) have identified common variants associated with hepatitis C virus (HCV)-induced liver disease. Two GWASs of fibrosis progression have been performed in HCV-infected Europeans and identified three loci near *RNF7*, *MERTK/TULP1* and *CAV3/RAD18*^11,12^. The only GWAS of HCV-induced cirrhosis, conducted in a Japanese population, identified two HLA alleles as risk factors^13^. For HCV-induced HCC, 4 GWASs in East Asian populations have identified loci near *MICA, DEPDC5*, the HLA region and *TLL1*^14,15,16,17^. A recent study in a North American population also found an HLA allele associated with HCV-induced HCC^18^. However, the replication of these risk factors has been inconsistent across studies, often due to challenges in establishing large and well-powered cohorts of infected patients with detailed clinical data. Furthermore, phenotype definitions, including the use of uninfected or unexposed population-based controls, can bias association results^19^. The interaction between risk variants and other viral or host risk factors also remains largely unexplored.

Early identification of at-risk patients is crucial for reducing liver mortality, yet current risk stratification tools are inadequate^20,21^. Polygenic risk scores (PRS) have been developed for a variety of liver-related traits and diseases in the context of non-viral aetiologies^22,23^, yet their validity in HCV-infected patients remains unclear.

In this study, we performed a GWAS of cirrhosis and HCC in a multi-ethnic cohort of HCV-infected patients recruited from routine hospital care across the United Kingdom and clinical trials. We examined the interaction of common genetic variants with other host and viral risk factors. We also analysed liver gene expression from infected patients and identified disease-specific expression quantitative-trait loci (eQTL) to link noncoding variants to putative effector genes, and test whether cirrhosis or cell-type composition modifies genetic effects. Finally, we examined the clinical utility of PRS derived from non-infected populations by testing them in our study cohort.

## Materials and Methods

### Patient subjects

The samples used for this case-control analysis were derived from the STOP-HCV consortium that combined three cohorts of patients with chronic HCV infection as previously described^24,25^. Briefly, the first cohort included patients recruited by the HCV Research UK (HCVRUK) clinical database and biobank that includes over 10,000 patients from across the United Kingdom, who have attended a specialist HCV clinic for care/management of their HCV infection between March 2012 and July 2015. These patients presented with no liver disease, a compensated liver disease, or advanced liver disease (decompensated cirrhosis) and a minority subsequently developed hepatocellular carcinoma (HCC). In particular, two nested studies were included within HCVRUK: 806 patients with advanced cirrhosis who were treated under the NHS England Expanded Access Programme (EAP) launched in 2014, giving early access to first-generation all-oral antivirals; and (ii) 1,255 patients with compensated cirrhosis, recruited from 31 hospital sites across the UK from January 2015 to September 2016, and prospectively followed in the STOP-HCV Cirrhosis Study, a 5-year investigation of progression to decompensation and HCC^26^. Cirrhosis was defined on the basis of: (i) histological assessment (Ishak 5/6 or Metavir 4); or (ii) imaging results consistent with cirrhosis, including Fibroscan >15 kPa; or (iii) validated serum biomarker consistent with cirrhosis (including APRI >2 and Enhanced Liver Fibrosis [ELF®] test >10.48). HCC was ascertained from clinician reports with reference to patient hospital records. For participants from England, long-term follow-up data were obtained by linkage to national records for hospital admission, cancer registrations, and mortality. A case was recorded when diagnosis was supported by one of the following: liver biopsy, transplant histology, cross-sectional imaging, tumour markers, imaging plus tumour markers, or post-mortem examination. The majority of the HCVRUK patients were infected with either genotype 1 or 3 HCV.

The second cohort included patients from the BOSON phase 3 randomized open-label trial who were recruited in five different countries (Australia, Canada, New Zealand, United Kingdom and United States) and had no or compensated cirrhosis^27,28^. Presence of cirrhosis was determined by liver biopsy or by a FibroScan result of > 12.5 kPa. The majority of the BOSON patients were infected with genotype 3 HCV. The third cohort consisted of British patients from the STOP-HCV-1 post-licensing randomised controlled trial which recruited individuals without evidence of liver fibrosis (FibroScan score ≤ 7.1 kPa)^29^. All STOP-HCV-1 patients were infected with genotype 1 HCV.

The sampling protocols for all cohorts were approved by the appropriate institutional review boards, and all patients provided written informed consent. All studies conform to the ethical guidelines of the 1975 Declaration of Helsinki. The HCVRUK patients were enrolled by consent into the HCV Research UK registry. Ethics approval for HCV Research UK was given by NRES Committee East Midlands – Derby 1 (Research Ethics Committee reference 11/EM/0314). The BOSON study protocol was approved by each institution’s review board or ethics committee before study initiation (clinical trial registration number: NCT01962441). The STOPHCV1 trial was approved by the Cambridgeshire South Research Ethics Committee (15/EE/0435). The trial was registered at ISRCTN (37915093, 11 th April 2016), and EudraCT (2015-005004-28, 31 st December 2015).

In total, 4,344 patients with available genetic and clinical data were further retained for the case-control analyses (523 from the BOSON cohort, 187 from the STOP-HCV-1 cohort and 3,634 from the HCVRUK cohort (including 1149 from cirrhosis sub cohort and 527 from EAP sub cohort)). The characteristics of these patients are presented in Table 1. We performed all subsequent statistical analyses in R (version 4.2.0)^30^.

**Table 1.** Demographic and clinical characteristics of the STOP-HCV GWAS patients.

|  | **HCVruk cohort** | **BOSON cohort** | **STOP-HCV-1 cohort** | **Total** |
| --- | --- | --- | --- | --- |
| n | 3,634 | 523 | 187 | 4,344 |
| Age at enrolment, years, mean (sd) | 51.9 (10.8) | 49.8 (10.0) | 45.7 (10.7) | 51.4 (10.8) |
| Sex, female, n (%) | 1,040 (28.6) | 178 (34.0) | 57 (30.5) | 1,275 (29.4) |
| BMI, mean (sd) | 27.3 (5.1) | 27.7 (5.3) | 25.3 (4.6) | 27.3 (5.1) |
| Type 2 diabetes, n (%) | 651 (18.3)<sup>a</sup> | 32 (6.1) | 1 (0.5) | 684 (16.0) |
| Alcohol misuse, n (%) | 1,562 (43.6)<sup>b</sup> | 19 (3.6) | 34 (18.2) | 1,615 (37.6) |
| HIV co-infection, n (%) | 135 (3.9)<sup>c</sup> | 0 (0) | 66 (35.3) | 201 (4.8) |
| HCV genotype, n (%)<sup>d</sup> |  |  |  |  |
| Genotype 1 | 1,069 (31.3) | 1 (0) | 184 (98) | 1,254 (30.7) |
| Genotype 3 | 2,345 (68.7) | 479 (100) | 0 (0) | 2,824 (69.2) |
| Cirrhosis (any), n (%) | 2,468 (67.9) | 361 (69.0) | 0 (0) | 2,829 (65.1) |
| Cirrhosis (decompensated), n (%) | 939 (25.8) | 0 (0) | 0 (0) | 939 (21.6) |
| Hepatocellular carcinoma, n (%) | 706 (19.4) | 0 (0) | 0 (0) | 706 (16.2) |
| Liver transplant, n (%) | 494 (13.6) | NA | NA | 494 (11.4) |
| Death, n (%) | 535 (14.7) | NA | NA | 535 (12.3) |

<sup>a</sup> Missing information for 69 patients<sup>b</sup> Missing information for 51 patients<sup>c</sup> Missing information for 187 patients<sup>d</sup> A total of 266 patients had either another HCV genotype or missing information

### Genotyping and imputation procedures

Blood specimens collected at enrolment in each cohort were used to generate host-genotyping data using the Affymetrix UK Biobank array, which covers ∼820,000 markers. Using PLINK v1.9^31^, we performed quality control by excluding single nucleotide polymorphisms (SNPs) with a minor allele frequency (MAF) < 5%, call rate of < 98 % and Hardy-Weinberg equilibrium *P* < 1 × 10^−5^. We also filtered out SNPs showing significant batch difference or sex difference in allele frequency (*P* < 1 × 10^−5^). This process resulted in a total of 542,730 high-quality autosomal SNPs retained for imputation.

Individuals were excluded if they had a call rate < 95% or an outlier heterozygosity value (±3 SD from the mean of their self-identified ancestry group). The sex of each subject was verified by comparing the reported sex with the observed sex based on X chromosome method-of-moments F coefficient. To identify duplicates, we assessed pairwise genotype concordance and performed an identity-by-descent (IBD) analysis. For pairs with genotype concordance > 95% at a set of pruned SNPs, the individual with the lower call rate was excluded.

Imputation was performed on the TOPMed Imputation Server using the TOPMed reference panel (version r2)^32^. Approximately 6.3 million SNPs with an imputation quality score greater than 0.3 were retained for genetic analyses. Classical HLA alleles and amino acid residues were imputed using SNP2HLA^33^ and the UK Biobank as the reference panel^34^ and only common alleles (MAF > 1%) with an imputation accuracy r² > 0.8 were kept for analyses.

### Principal component analysis

We conducted a principal component analysis (PCA) to evaluate population substructure. Kinship coefficients were estimated using PC-Relate (as implemented in the R package GENESIS)^35^ to select unrelated individuals. The R package SNPRelate^36^ was then used for PCA on pruned common genotyped SNPs from a set of unrelated individuals, defined as those with pairwise kinship coefficients less than 2^-9/2^. These two estimation procedures were iterated to ensure that the kinship coefficients were unbiased in admixed individuals and that the PCs were computed on an unrelated set. PC scores were then estimated for all remaining individuals by projection (Supplementary Figure 1). To define a European subgroup, ADMIXTURE^37^ was used with K = 5 to infer continental ancestry fractions, using reference populations from 1000 Genomes Project^38^. Individuals were attributed to a European ancestry group when their European ancestry percentage was > 80% (Supplementary Figure 2).

### Genetic analyses

Association testing between each common imputed SNP and liver disease phenotypes was performed on each autosome using an additive generalized linear mixed model, as implemented in the R package GENESIS. To account for genetic ancestry and relatedness, we included a genetic relatedness matrix (GRM) as a random effect in the model, computed on the genotyped data and for each chromosome separately in a leave-one-chromosome-out (LOCO) fashion. Age at enrolment, sex and the first 10 PCs were included as fixed covariates. The top 10 PCs which captured most of the genetic variation in the cohort were included to account for population structure (Supplementary Figure 3). Quantile–quantile plots and genomic control factor λ were used to assess genomic inflation.

For the HLA fine-mapping, we tested for association with liver disease using an additive logistic regression model with the same covariates as in the GWAS. We considered HLA alleles at two-field resolution, as well as biallelic and multiallelic amino acid variants. To identify independent signals, we performed iterative conditional analyses by adding the most strongly associated HLA allele or amino acid position to the model in each step. The p-value threshold significance was set at 4.80×10^−4^ for a total of 104 HLA alleles tested and 1.42 ×10⁻^4^ for a total of 353 amino acid positions tested. To identify the most strongly associated residue at each position, we used the alignment provided by the IMGT-HLA^39^ to determine residue positions and we performed an omnibus test by comparing a full model (including M-1 amino acid dosages) to a reduced model via an F-test on the delta deviance. Protein structures of the HLA-DQ and HLA-DR molecules were downloaded from the Protein Data Bank (PDB)^40^ and the location of amino acid residues on HLA proteins was visualized with PyMol v.3.1 (The PyMOL Molecular Graphics System, Version 3.0 Schrödinger, LLC).

We investigated two primary phenotypes: cirrhosis and HCC. For cirrhosis, patients were defined as cases with any diagnosis of cirrhosis at any time between enrolment and the end of follow-up in August 2020, and controls were chronically infected patients with no evidence of cirrhosis. For HCC, patients were defined as cases with any diagnosis of HCC at any time during the same period (with or without cirrhosis) and controls were cirrhosis patients only. For interaction analyses, the cohort was further stratified by virus genotypes (gt1 vs. gt3), alcohol misuse (yes/no), BMI > 25 kg/m² and type 2 diabetes (or BMI < 25 and absence of T2D). Alcohol misuse was defined as a history of, or current, heavy drinking with consumption of > 14 units per week for 6 months or more and/or use of medication for alcohol use disorder. The association with end-stage liver disease (decompensated cirrhosis and liver transplant) and all-cause mortality (any recorded death between enrolment and the end of follow-up in August 2020) was also examined. To assess the significance of the interaction between host SNPs and virus genotypes (genotype-by-environment interaction), a Wald test for the interaction term in the regression was used as implemented in the R package GENESIS, considering *P* < 0.05 as statistically significant.

### eQTL analysis

Liver biopsy samples were available for 194 patients infected with HCV genotype 3 from the BOSON trial. RNA extraction, library preparation and sequencing have been described elsewhere^41,42^. Read pairs were mapped to human genome GRCh38 using STAR aligner v.2.7.10b^43^. Uniquely mapped read pairs were counted using featureCounts v.2.0.3^44^ and GENCODE v.42 for gene annotation. Four samples were excluded due to low sequencing depth, which left 190 samples with available genetic data for analysis, including 54 with cirrhosis.

Low-expressed genes were filtered out when they had a count per million (CPM) > 0 in at least half of the samples using the R package edgeR^45^. RPKM values (Reads Per Kilobase Million) were calculated after normalization by effective library sizes with the trimmed mean of M values (TMM) method and by gene length and were transformed to TPM values (Transcripts Per Million). The log_2_(TPM+1) values were used in the eQTL analysis, which included 17,302 protein-coding and noncoding RNA genes. We performed PCA on the normalized gene counts matrix with the R package prcomp (center = true, scale = true) (Supplementary Figure 4).

We performed association testing between each common imputed SNP under an additive model and normalized gene expressions in ±1 Mb *cis* window centred around the start of each gene, using a linear regression as implemented in the R package Matrix eQTL^46^. We investigated the association between the gene expression PCs and a set of clinical variables that may influence gene expression and observed that the first 10 PCs captured most of the expression variation (Supplementary Figure 5). We included these 10 RNA PCs, the 5 first genetic PCs and sex as covariates. To identify SNPs with different effect sizes on gene expression in presence of cirrhosis (cirrhosis interaction *cis*-eQTLs, cirrhosis ieQTLs), we added cirrhosis status and a cirrhosis-SNP interaction term to the model. Multiple hypothesis testing correction for *cis*-QTLs was carried out using EigenMT^47^, which estimates the number of effective tests for each gene by considering the LD between SNPs. Then, *q*-values were calculated after adjusting the obtained EigenMT *P* values across the genes. We defined as eGenes and cirrhosis ieGenes the genes harbouring a significant eQTL with *q* < 0.05.

We conducted a colocalization analysis using the coloc method^48^ with default parameters to test whether the two association signals from the GWAS and the eQTL analysis were consistent with a shared causal variant. Then, we estimated liver cell-type abundances in the BOSON patients and GTEx samples^49^ (v8) using the DWLS method as implemented in the R package omnideconv^50,51^. Gene expression signature matrices were derived from 2 scRNA-seq atlases: a dataset of 5 healthy liver samples with 8,444 cells clustered into 20 cell populations(MacParland et al^52^) and a dataset of 5 healthy and 5 cirrhotic samples with 66,135 cells clustered into 12 cell lineages (Ramachandran et al^53^). For each atlas, genes differentially expressed across annotated cell types were compiled into a signature matrix, which is used as input to estimate relative cell-type proportions in the BOSON and GTEx bulk liver RNA-seq samples. Cell-type compositions were compared using pairwise two-sample Wilcoxon tests between non-infected GTEx samples, HCV-infected BOSON patients with normal and cirrhotic liver. *P* values were Bonferroni-corrected. Finally, to identify cis-eQTLs whose effects are dependent on cellular context (cell-type interaction eQTLs, or ieQTLs), we performed a separate analysis for each of the 13 most abundant cell types or lineages (average proportion > 1%). For each cell type, we used a linear model that included the cell-type proportion as a variable and an interaction term between genotype and this proportion. The significance of this interaction term was tested to identify variants whose effect on gene expression is modified by the abundance of that specific cell type. Multiple testing was controlled with EigenMT, followed by *q*-value estimation and further Bonferroni correction for the 13 cell types. Genes harbouring at least one eQTL with *q* < 0.0038 (0.05/13) were designated cell-type ieGenes.

### Polygenic risk scores testing

Polygenic risk scores (PRS) of cirrhosis, alcohol-related liver disease (ARLD) and non-alcoholic fatty liver disease (NAFLD) were downloaded from the Polygenic Score Catalog^54,55^. Details of the 10 PRS are reported in Supplementary Table 1. The pgs_calc tool was then used to calculate scores for the European subgroup of the cohort. PRS were Z-score normalized and centred (mean = 0, standard deviation = 1) to facilitate interpretation of odds ratios (OR), that is per standard deviation (SD) of the PRS. We used logistic regression to test for association between the PRS and cirrhosis or HCC status correcting for age, sex, and the first 10 genetic PCs. Interaction analyses were conducted similarly to the GWAS. We calculated the area under the curve (AUC) using the R package pROC^56^.

## Results

### Genome-wide association and gene-virus interaction study of cirrhosis and HCC

First, we conducted a GWAS of 2,829 HCV-induced cirrhosis cases and 1,515 HCV-infected controls without evidence of cirrhosis (Figure 1). We tested a total of 6,504,702 variants and observed a genomic inflation factor (λ) of 1.008, indicating an adequate control of population structure and relatedness (Supplementary Figure 6). We observed a genome-wide significant association on chromosome 22, corresponding to a cluster of three variants in *PNPLA3.* The cluster comprised the missense variant rs738409 C>G (p.I148M), associated with an odds ratio (OR) per G allele of 1.40 (95% confidence interval [CI] = 1.25–1.57, *P* = 1.03×10⁻^8^) (Figure 2A).

**Figure 1.**
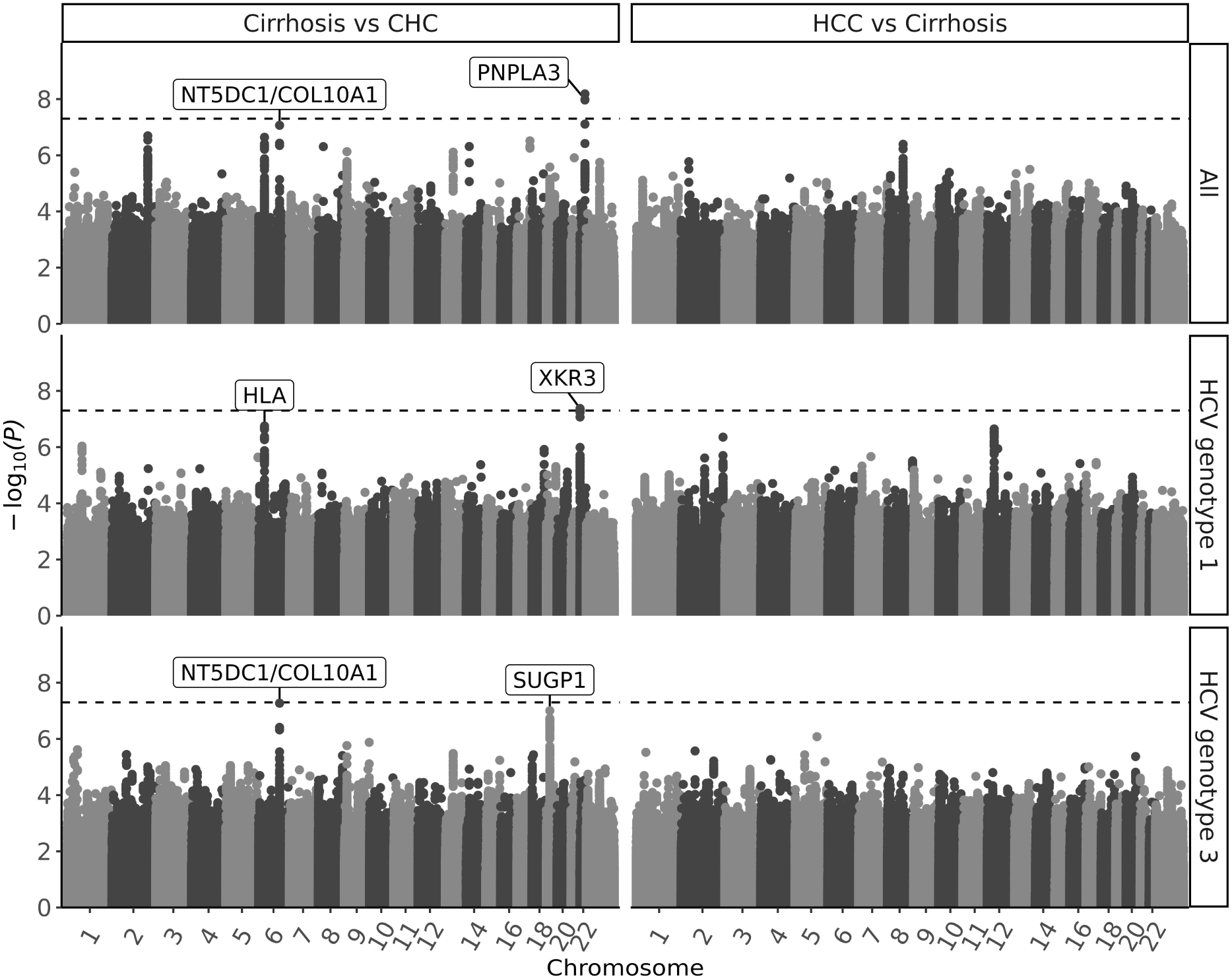
GWAS of liver disease in the STOP-HCV cohort. Manhattan plots showing the – log_10_(p-value) of the association of common genetic variants with HCV-induced cirrhosis, using chronic hepatitis C (CHC) as control group (left panel) and with HCV-induced hepatocellular carcinoma (HCC) using cirrhosis as control group (right panel), in all patients and stratified by virus genotype. The dashed line corresponds to the genome-wide significance threshold.

**Figure 2.**
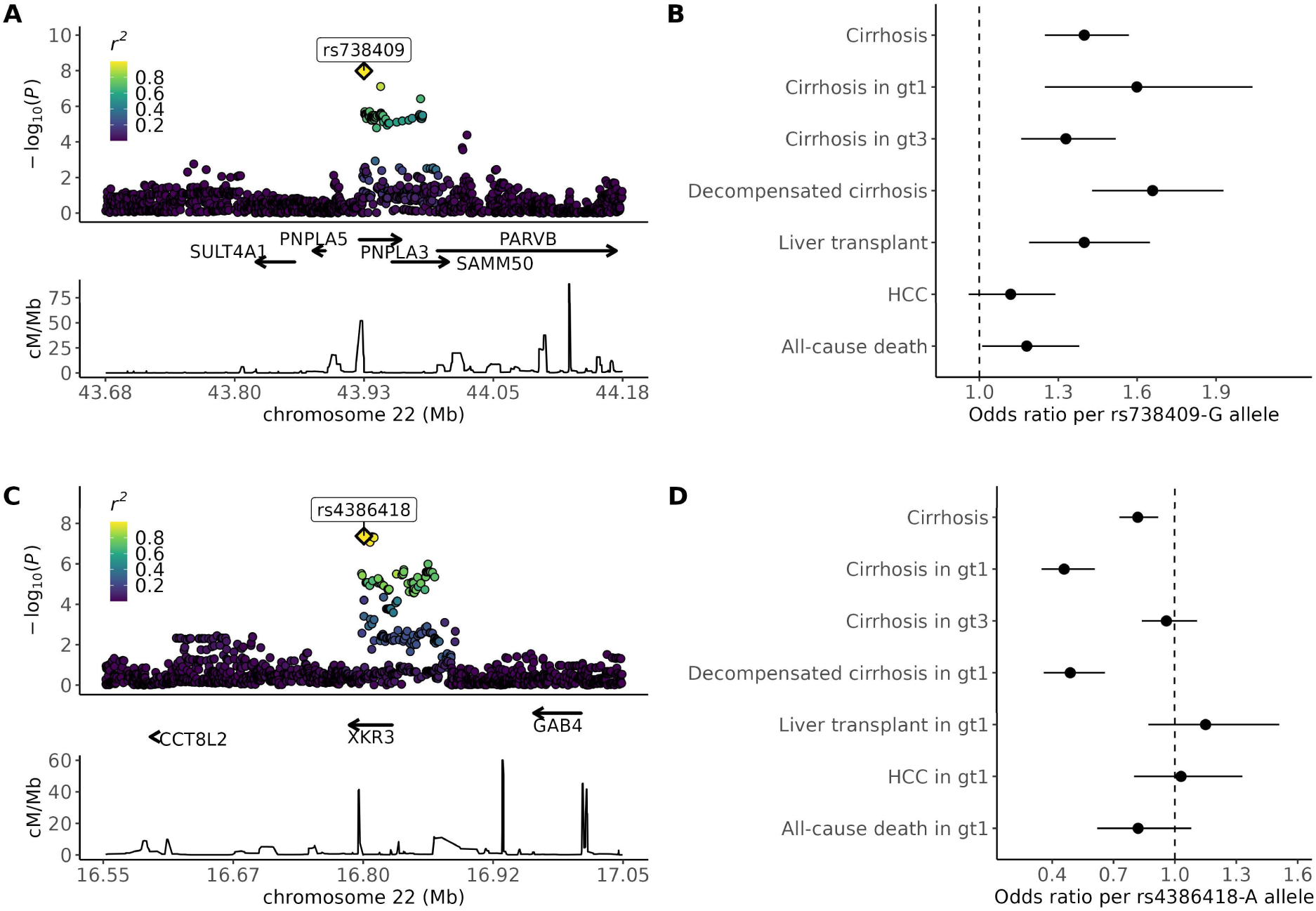
Regional association plots and forest plots for the two genome-wide significant loci. **A)** Regional plot for the *PNPLA3* risk locus. Each dot represents the –log10(p-value) of the association of a common variant with cirrhosis risk. The second most significant associated variant (the missense rs738409 or p.I148M) is represented by the yellow square. The colour of all other variants indicates the level of linkage disequilibrium with rs738409 (using 1000 Genomes Project populations of European ancestry). Recombination rates were also estimated from 1000G. **B**) Forest plot for the rs738409 variant showing odds ratios and 95% confidence intervals for liver disease and death. **C)** Regional plot for the *XKR3* locus identified in patients infected with HCV genotype 1. **D)** Forest plot for the rs4386418 variant.

Then, we conducted a GWAS of 706 HCV-induced HCC cases (including 677 cases with cirrhosis and 29 cases who developed HCC without evidence of cirrhosis) and 2,152 cirrhosis controls. No genome-wide significant association was observed (Figure 1), even after adding 1,486 infected patients without evidence of cirrhosis to the control group (λ = 1.00 for both analyses) (Supplementary Figure 7). The *PNPLA3* rs738409 risk allele was not associated with the risk of HCC in cirrhosis patients (OR = 1.12, 95% CI = 0.96–1.29, *P* = 0.14) (Figure 2B). In addition, none of the loci previously reported to be associated with HCC (induced by HCV or other causes) replicated, except the HSD17B13 rs72613567 variant (Supplementary Figure 8).

Next, we stratified the GWAS of cirrhosis and HCC by virus genotype to investigate potential host-virus interactions. We restricted this analysis to HCV genotypes 1 and 3 for which the sample sizes were sufficient (gt1: 1,020 cirrhosis and 292 HCC cases for 234 and 738 controls, respectively; gt3: 1,555 cirrhosis and 348 HCC cases for 1,270 and 2,477 controls, respectively). The *PNPLA3* rs738409 risk allele was associated with cirrhosis, in both virus genotypes (*P* = 2.01×10^−4^ and 5.07×10⁻^5^ in gt1 and gt3, respectively, *P* for interaction = 0.40). We observed a genome-wide significant association with protection against cirrhosis in gt1-infected patients on chromosome 22, corresponding to a cluster of five intronic variants in *XKR3* with the lead variant being rs4386418 (OR per A allele = 0.46, 95%CI = 0.35–0.61, *P* = 4.22×10⁻^8^) (Figure 1, Figure 2C). This association was not found in gt3 patients (OR per A allele = 0.96, 95%CI = 0.84–1.11, *P* = 0.60), and the interaction was significant (*P* for interaction = 5.37×10⁻^6^, Figure 2D). The rs4386418 protective allele was not associated with HCC in gt1 cirrhosis patients (OR = 1.03, 95%CI = 0.80–1.33, *P* = 0.80).

In addition, we identified two loci with suggestive significance in gt3 patients (Supplementary Figure 9). First, an intronic variant in *NT5DC1*, next to *COL10A*, showed an association close to genome-wide significance (rs7755367, OR per T allele = 0.60, 95%CI = 0.50–0.72, *P* = 5.34×10⁻^8^). While not significant in gt1 patients, the effect size was in the same direction and no interaction was observed (OR = 0.89, *P* = 0.13 in gt1, *P* for interaction = 0.14) (Supplementary Table 2). Second, a large cluster of variants on chromosome 19p13 had the strongest association for an indel located in *SUGP1* (rs148030853, OR per del = 1.38, 95%CI = 1.22–1.55, *P* = 9.89×10⁻^8^). This association was not found in gt1 patients, as confirmed by a significant interaction (*P* for interaction = 0.01).

We looked at the association of the *PNPLA3* rs738409 risk variant and the gt1-specific *XKR3* rs4386418 protective variant with end-stage liver and all-cause mortality (Supplementary Tables 3 and 4). Both variants increased the risk of decompensated cirrhosis (*P* = 6.90×10⁻^11^ and 6.16×10⁻^6^, respectively) but only the rs738409 risk variant was associated with liver transplant (*P* = 5.47×10⁻^5^ and 0.33, respectively). In addition, the rs738409 risk variant was also associated with all-cause mortality (*P* = 0.03). The effect sizes of both variants were consistent across strata of alcohol misuse, BMI, and type 2 diabetes.

### HLA fine-mapping

Previous studies identified *HLA-DQA1*06:01* as a risk factor for HCV-induced cirrhosis^13^ and *HLA-DQB1*03:01* and *DQB1*06:02* for HCC^16,18^. In the present GWAS of cirrhosis, we observed a signal in the MHC region at a suggestive significance threshold (rs17212223, OR per T allele = 0.69, 95%CI = 0.60–0.80, *P* = 2.29×10⁻^7^). We then imputed classical HLA alleles and tested their association with liver disease and their interaction with virus genotypes. We observed four significant associations with cirrhosis *HLA-DQB1*03:01*, (*P* = 1.58×10⁻^7^), *HLA-DRB1*13:01* (*P* = 2.82×10⁻^5^), *HLA-DQA1*01:03* (*P* = 5.69×10⁻^7^) and *HLA-DQB1*06:03* (*P* = 8.08×10⁻^5^) (Supplementary Table 5). In a conditional analysis, *HLA-DQB1*03:01* and *HLA-DRB1*13:01* were independently associated with cirrhosis and including them in the model removed all residual associations (Supplementary Table 6). *HLA-DQB1*03:01* had a protective effect against cirrhosis (OR = 0.69, 95%CI = 0.59–0.79) while *HLA-DRB1*13:01* increased the risk (OR = 1.53, 95%CI = 1.25–1.86). Both alleles showed consistent direction of effects in gt1 and gt3 patients, but estimates were larger in gt1, with a significant interaction for *HLA-DQB1*03:01* (*P* for interaction = 0.001) (Supplementary Table 7). None of the alleles were associated with progression to HCC (Supplementary Table 8).

We next identified specific amino acid residues in the HLA-DQβ1 and HLA-DRβ1 proteins associated with cirrhosis. The strongest association was observed for the multiallelic position 37 in HLA-DRβ1 (*P* = 2.00×10⁻^8^) (Supplementary Table 9). The omnibus test showed that carrying an Asparagine (Asn) at this position was associated with an increased risk of cirrhosis (OR = 1.25, 95% CI = 1.10–1.52, *P* = 4.81×10⁻^4^) while carrying a Tyrosine (Tyr) had a protective effect (OR = 0.84, 95% CI = 0.74–0.95, *P* = 5.47×10⁻^3^) (Supplementary Table 10). After conditioning on HLA-DRβ1 Asn37, we observed a signal at position 45 of HLA-DQβ1 (*P* = 1.20×10⁻^5^) (Supplementary Table 11). This position corresponded to a biallelic amino acid variant (Glutamic Acid/Glycine) where Glu drove the association (OR = 0.73, 95% CI = 0.63– 0.84, *P* = 1.64×10⁻^5^). No further associations were found after conditioning on both amino acids, suggesting that these two amino acids drove the association with cirrhosis (Supplementary Table 11). HLA-DRβ1 Asn37 is in the peptide binding pocket 9 of the β chain of HLA-DR. HLA-DQβ1 Glu45 is in the β-turn of the HLA-DQ protein, which connects the α-helix and β-sheet and maintains the conformational structure of the antigen binding groove (Supplementary Figure 10).

### Cirrhosis-interaction and cell-type-interaction eQTL mapping in HCV-infected liver

To investigate the regulatory mechanisms of gene expression in HCV infection and cirrhosis, we performed a *cis*-eQTL analysis for 190 infected liver samples from the BOSON trial, including 54 cirrhosis cases. In total, we tested 6,314,313 variants and 17,302 genes. We identified 398,933 significant SNP-gene pairs, corresponding to 2,060 genes with eQTLs (eGenes, *q* < 0.05) (Supplementary Table 12). Next, we performed an interaction analysis to examine cirrhosis-specific effects. We identified 2,396 significant SNP-gene pairs, corresponding to 129 genes with cirrhosis-specific effect sizes (ieGenes, Supplementary Table 13). We compared BOSON (HCV-infected liver) eGenes to those from GTEx (healthy liver) (Figure 3A and B) and found that 24.4% of BOSON eGenes (1,403/5,734) overlapped with GTEx liver eGenes, the second largest overlap across all 48 GTEx tissues tested (Supplementary Figure 11). BOSON eGenes displayed higher expression than non-eGenes in both BOSON and GTEx datasets (Supplementary Figure 12). Furthermore, BOSON eQTLs unique to this study (not found in GTEx) showed smaller effect size correlations with GTEx data compared with eQTLs shared between both datasets, suggesting these may represent regulatory effects specific to chronic HCV infection. However, this pattern did not apply to cirrhosis interaction eQTLs (Supplementary Figure 13).

**Figure 3.**
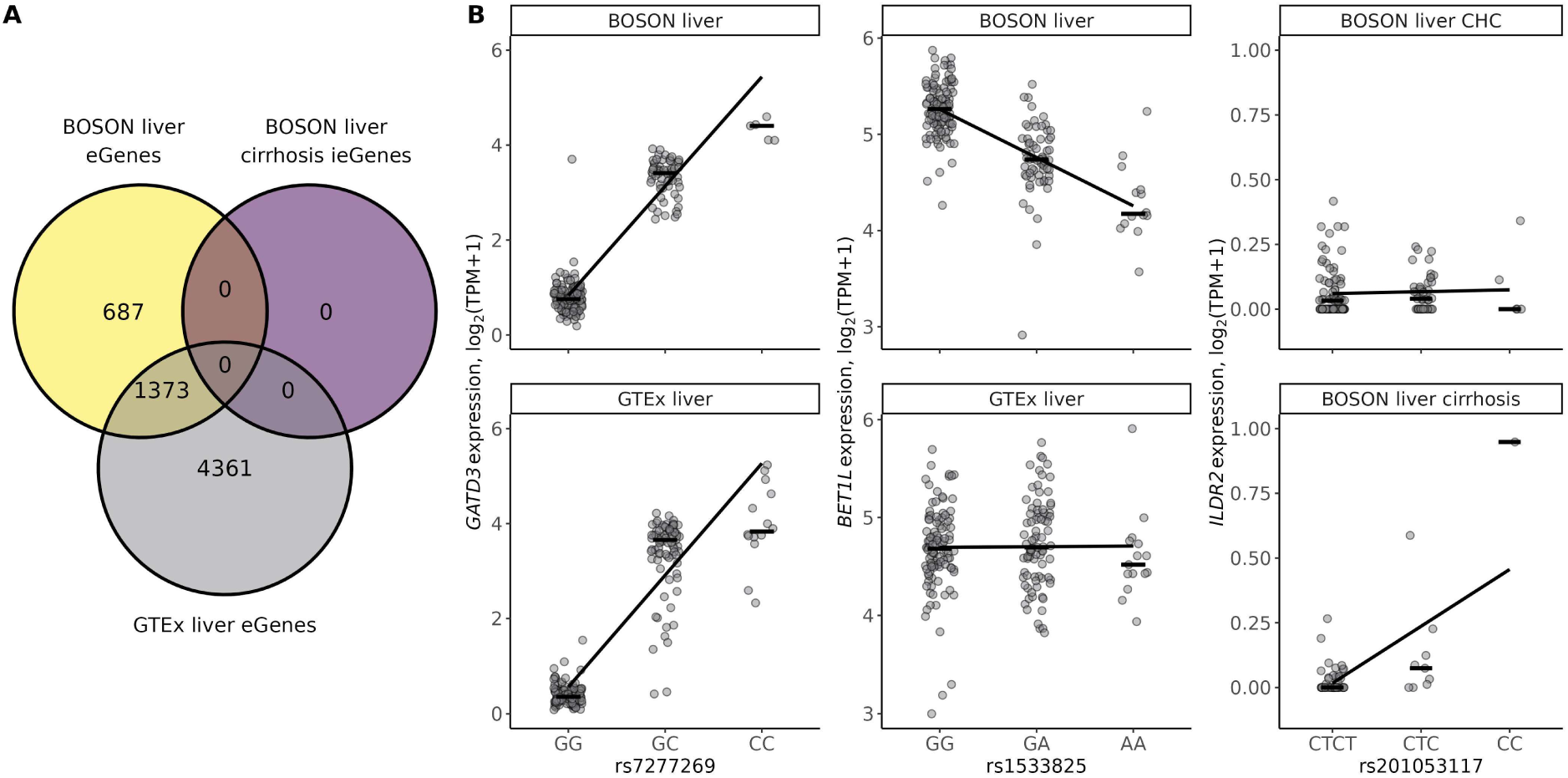
*Cis*-eQTL mapping in HCV infected liver biopsies from BOSON trial. **A)** Overlap of genes with eQTLs (eGenes) from BOSON and genes with cirrhosis specific eQTLs (ieGenes) with GTEx v8 liver (healthy) eGenes. **B)** Examples of eQTLs shared between BOSON and GTEx (left panel), specific to BOSON (middle panel) and specific to cirrhosis in BOSON (right panel).

Next, we examined the genetic regulation of *PNPLA3* expression in the liver and its relationship to cirrhosis risk in CHC. *PNPLA3* expression did not differ significantly between CHC and cirrhosis patients (Figure 4A). We identified an intronic variant (rs4823174) as a significant eQTL associated with a decreased expression of *PNPLA3* (beta per A allele = –0.20, *P* = 1.57×10⁻^6^). However, this variant is in low LD (r² < 0.2) with the rs738409 GWAS risk variant and was not associated with the risk of cirrhosis in the GWAS (*P* = 0.09) (Figure 4B and C). Conversely, the rs738409 risk variant was not associated with *PNPLA3* expression (beta per G allele = 0.03, *P* = 0.42). The colocalization analysis suggested that PNPLA3 expression and cirrhosis risk are likely influenced by different variants and there is a limited support for a shared causal variant between GWAS and eQTL signal (H3 = 0.49, H4 = 0.006).

**Figure 4.**
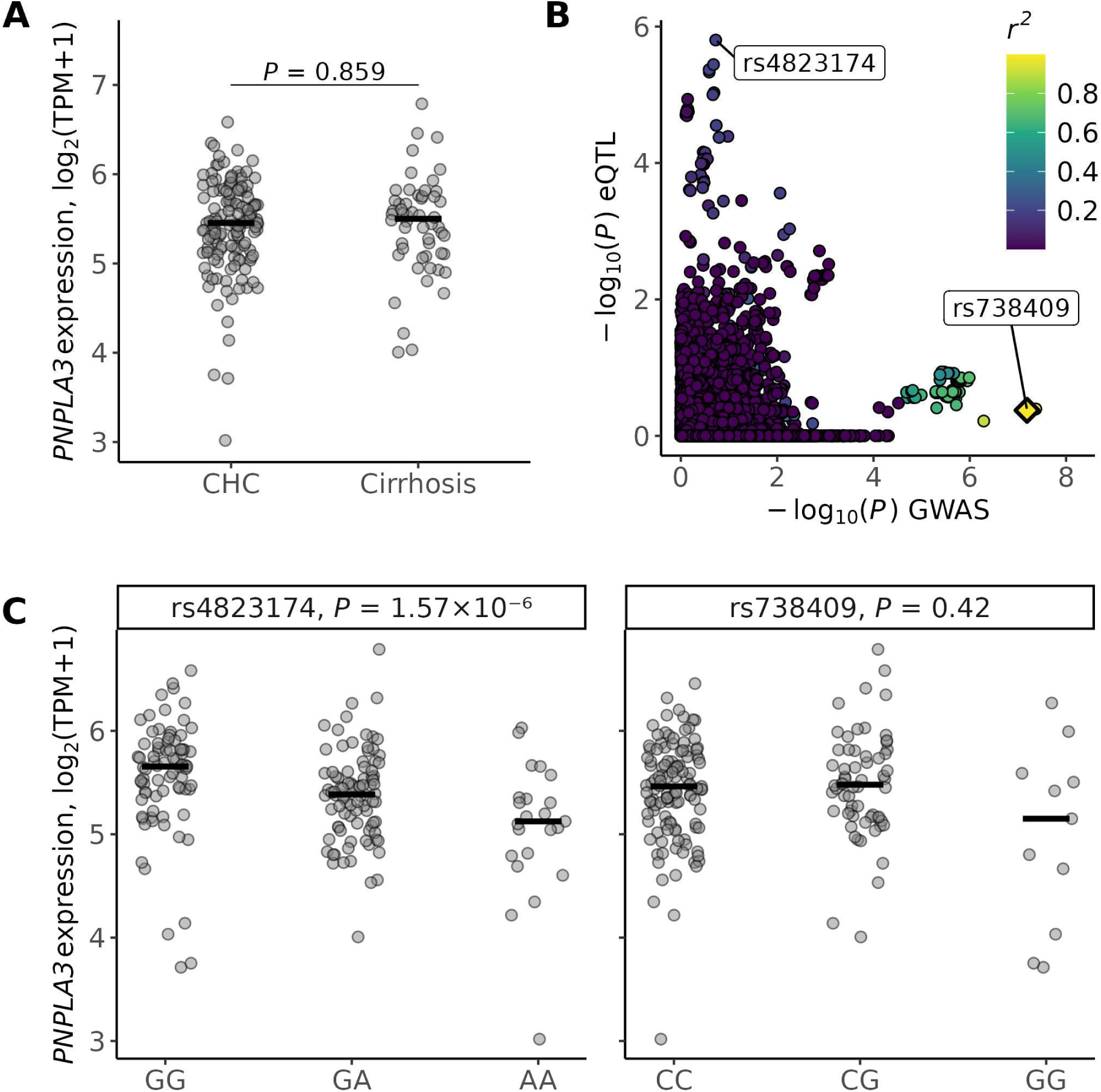
eQTL mapping for *PNPLA3* in HCV-infected liver from the BOSON trial. **A)** *PNPLA3* liver expression by cirrhosis status. **B)** Scatter plot showing the overlap of the GWAS and eQTL results in the *PNPLA3* locus. The x-axis represents the –log10(p-values) of the GWAS for cirrhosis and the y-axis represents the –log10(p-values) of the eQTL mapping for *PNPLA3*. The degree of linkage disequilibrium for all variants with rs738409 is indicated by colour. **C)** Boxplots showing *PNPLA3* expression levels for the genotypes of the top eQTL variant rs4823174 and the top GWAS variant rs738409.

Next, using deconvolution, we estimated cell-type proportions in the BOSON and GTEx bulk liver RNA-seq samples, employing signature matrices constructed from the MacParland healthy liver atlas and the Ramachandran atlas, which includes both healthy and cirrhotic samples (Supplementary Figure 14 and Table 14).

Using the MacParland atlas, certain hepatocyte clusters (hepatocytes_4 and hepatocytes_3) were dominant in healthy GTEx liver comprising 49% and 33% of cells, respectively. In BOSON patients with chronic hepatitis C (CHC), these proportions dropped significantly to 34 % (*P* = 7.41×10⁻⁹) and 2.5 % (*P* = 1.85×10⁻²⁹) and remained similarly low in cirrhosis (26 %, *P* = 1.58×10⁻⁸; 6.1 %, *P* = 9.42×10⁻¹¹). The differences between CHC and cirrhosis was not significant for either cluster (Supplementary Table 15).

In contrast, Plasma B cells, mature B cells and portal endothelial cells were substantially increased in CHC (0.66%, 0.62%, and 0.78%, respectively) compared to GTEx (0.20%, 0.09%, and 0.05%; all *P* < 1×10⁻¹³). These cell types further expanded in cirrhosis, reaching 3.0%, 1.2%, and 1.2%, respectively (all CHC vs cirrhosis *P* < 1×10⁻³). Non-inflammatory macrophages were also enriched in CHC and cirrhosis (both 4.9 %) compared with GTEx (0.29 %, *P* = 6.42×10⁻⁵⁶ and *P* = 2.06×10⁻²⁹, respectively).

Using the Ramachandran atlas, we observed a similar trend. Hepatocytes declined from 80% in GTEx to 78% in CHC (*P* = 1.29×10⁻^4^) and further to 71% in cirrhosis (CHC vs cirrhosis, *P* = 3.60×10⁻^8^). Plasma cells rose sharply from 0.04% in GTEx to 8.0% (*P* = 2.06×10⁻⁵⁷) in CHC and to 12.5% in cirrhosis (CHC vs cirrhosis, *P* = 2.28×10⁻⁵). Mononuclear phagocytes and endothelial cells increased (6.3% and 3.4%) compared to GTEx (1.6% and 2.4%; *P* = 1.56×10⁻⁴² and *P* = 1.48×10⁻¹⁰), with similar levels observed in cirrhosis (6.4% and 3.1%; no significant difference from CHC). The mesenchymal compartment was depleted in both CHC (2.8%) and cirrhosis (5.1%) compared to GTEx (10.3%; P = 7.89×10⁻³⁹ and P = 7.98×10⁻⁷, respectively).

Finally, we assessed how cell-type composition influences eQTLs effects by identifying cell-type interacting eQTLs. We focused on the 13 cell types or lineages with an average proportion > 1%. We identified between 7 and 1,089 significant cell-type interaction eGenes corresponding to a total of 3,534 associations (*q* < 0.0038) and to 2,525 unique ieGenes with a significant association in at least one of the cell types (Supplementary Tables 16 and 17). Notably, there was no correlation between the abundance of a cell type and the number of ieGenes identified. Most ieGenes (n = 1,887) were specific to a single cell type. Additionally, 92% (n = 2,317) of these ieGenes were not identified in the bulk eQTL analysis, highlighting the additional resolution provided by cell-type-specific models. (Supplementary Table 17). For the subset of eQTLs detected in both cell-type interacting and bulk analyses, effect size correlations were moderate and like those observed for ieQTLs found only in the interaction model (Supplementary Figure 15).

### Polygenic risk score analysis

The applicability of PRS derived from non-viral liver disease cohorts, such as those for ARLD or NAFLD to HCV-infected populations remains uncertain. To evaluate this, we tested 10 PRS from the PGS Catalog, including 2 for all-cause cirrhosis, 4 for ARLD, and 4 for NAFLD, in up to 3,406 HCV-infected patients of European ancestry (Supplementary Table 1).

The PRS were constructed using between 3 and 1,089,806 variants, most of which were successfully genotyped or imputed in the STOP-HCV cohort (variant missingness ranged from 0–5%, except for one 8-SNP score with 12% missingness but only 1 variant missing). Correlations between the 10 PRS varied widely (Pearson r = 0.09–0.82), with the 183k-SNP score PGS000704^57^ showing the lowest correlation with the others (r = 0.09–0.16; Figure 5A).

**Figure 5.**
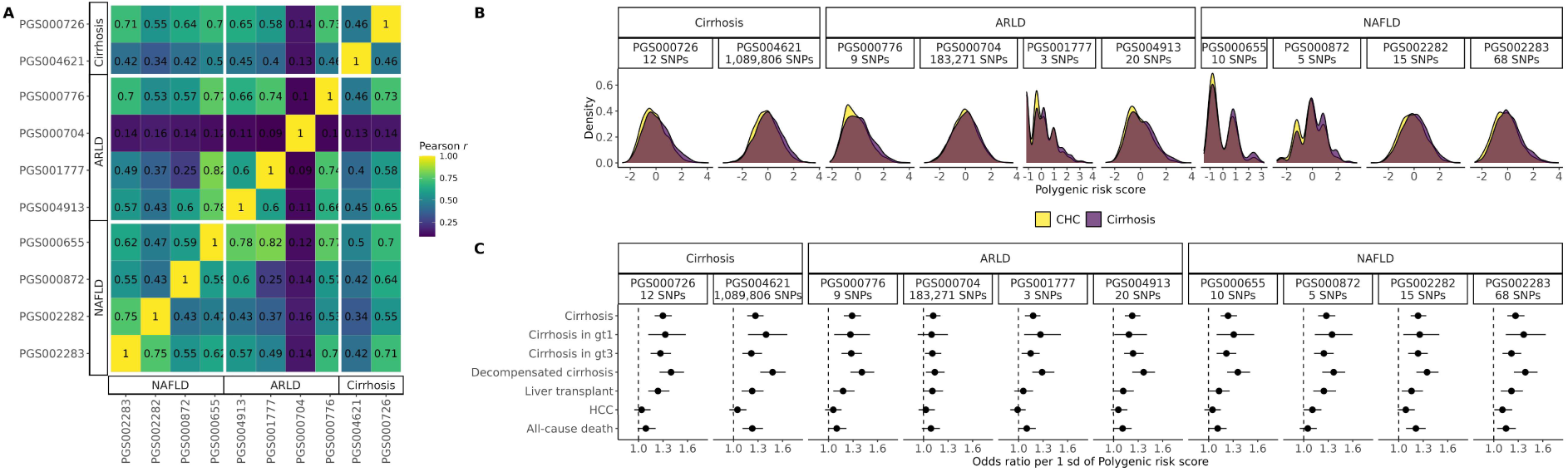
Validation of polygenic risk scores (PRS) of liver disease in the STOP-HCV cohort. **A)** Pairwise correlation between 10 normalized PRS. **B)** Distribution of the normalized PRS in the STOP-HCV cohort, stratified by cirrhosis status. **C)** Forest plot for the10 PRS showing odds ratios and 95% confidence intervals for liver disease and death.

All PRS showed higher average values in patients with HCV-induced cirrhosis compared to CHC controls (mean range: 0.03–0.08 in cases vs. –0.16–0.05 in controls; Figure 5B), and one standard deviation increase in PRS was consistently associated with increased odds of cirrhosis, ranging from 1.12 to 1.30 (Figure 5C). The strongest association was observed for a 12-SNPs score PGS000726, originally constructed for all-cause cirrhosis^23^ (OR = 1.30, 95%CI = 1.20–1.41, *P* = 2.10×10^−10^). The weakest association was observed for the 183k-SNPs PRS (PGS000704), which was based on a weighted combination of multiple PRS for ARLD biomarkers^57^ (OR = 1.12, 95%CI = 1.03–1.21, P = 5.71×10^−3^). The AUC for both PRS was moderate at 0.56 and 0.52, respectively (Supplementary Table 18).

We found no significant interaction between any PRS and virus genotype, alcohol misuse or diabetes/BMI status (Supplementary Table 18). However, all PRS showed a stronger association with decompensated cirrhosis with ORs ranging from 1.14 to 1.48. No PRS was significantly associated with HCC risk among cirrhosis patients, except for PGS002283, a 68-SNPs score originally constructed for NAFLD^58^, which showed a modest effect (OR = 1.11, 95%CI = 1.01–1.23, *P* = 0.03).

## Discussion

This study presents the first comprehensive GWAS and host genetic-virus genotype interaction analysis for cirrhosis and HCC, providing new insights into the genetic determinants of liver disease in the context of CHC. Our liver eQTL mapping identified HCV-infection and cirrhosis-specific eQTLs and in silico deconvolution highlighted cellular heterogeneity between healthy, HCV-infected, and cirrhotic liver. Furthermore, we showed that polygenic scores for non-viral liver diseases were associated with HCV-induced cirrhosis but not HCC. Our findings indicate that while the genetic architecture of HCV-induced cirrhosis overlaps with that of other aetiologies, the genetic drivers of HCV-related HCC are distinct from both HCV-associated cirrhosis and HCC in non-viral liver disease.

First, consistent with its well-known association with liver fat, NAFLD^59^ and ARLD^60^, the *PNPLA3* p.I148M variant also increased cirrhosis risk in our HCV infected cohort, whereas an independent eQTL that lowered *PNPLA3* mRNA showed no association with disease. This supports the view that the amino-acid change, rather than expression level, drives pathology in humans^61^. In addition, its effect remained independent from alcohol misuse, or type 2 diabetes and BMI. We also confirmed the lack of association between this variant and HCC development in the context of CHC, which contrasts with other etiologies^62,63,64^. Functionally, PNPLA3 p.I148M impairs lipid-droplet remodelling and regulates hepatic stellate-cell retinol metabolism, thereby amplifying steatosis and fibrogenic signalling, which are mechanisms that explain its consistent association with cirrhosis. The mechanisms by which p.I148M contributes to hepatocarcinogenesis remain unclear. The progression from fibrosis to cancer seems to require a persistent steatotic and inflammatory stress, which is more prominent in metabolic and alcohol-related liver disease^65,66^. In CHC, oncogenesis is unlikely to result from direct viral effects, as HCV does not integrate into the host genome. Thus, a sustained chronically inflamed and pro-tumorigenic environment is likely required to promote cancer development^67^.

Second, the observation of associations between *HLA-DQB1*03:01* and *HLA-DRB1*13:01* with cirrhosis risk is consistent with earlier research identifying HLA alleles as important determinants in the natural history of CHC. Notably, *HLA-DQB1*03:01* has been found to be associated with HCV spontaneous clearance, although our fine-mapping indicates that different amino acid residues are responsible for the cirrhosis association versus clearance^68,69^. Neither allele showed any association with HCC, contrary to previous studies reporting *HLA-DQB1* as a risk factor^16,18^. Interestingly, a Taiwanese GWAS found that the relationship between *HLA-DQB1* alleles and HCC risk varied according to HCV genotype^16^. This discrepancy may be due to differences in control group definitions^14,15,16,18^. Other studies often used non-HCC controls who may not have had cirrhosis (HCC very often happens in the context of cirrhosis), meaning their results likely captured a cirrhosis effect rather than a specific HCC effect. We mitigated this by using individuals with cirrhosis as control group.

This study also highlights the significance of host-virus interactions. The novel finding that the *XKR3* rs4386418 variant is associated with a reduced risk of cirrhosis specifically in gt1 patients warrants further investigation. This variant is an eQTL for *XKR3*, associated with increased expression in whole blood^70^. The function of the XKR3 protein is uncertain but it’s a close homolog of *XK*, which is a component of the Kell blood group complex and whose loss causes acanthocytosis in McLeod syndrome, suggesting a conserved role in membrane-lipid transport and cell-shape integrity^71^.

The eQTL analysis conducted in this study provides insights into the regulatory mechanisms of gene expression in CHC and cirrhosis. The overlap between eGenes from the BOSON trial and GTEx suggests that much of the genetic regulation of liver function may be conserved in CHC. Our ability to integrate GWAS and transcriptomic data was constrained by power: apart from PNPLA3 (and the genotype-1 XKR3 signal), we observed no additional genome-wide significant loci for cirrhosis or HCC, preventing us to systematically test for shared causal variants. In addition, because BOSON RNA-seq data came solely from gt3-infected patients, we were unable to assess the impact of viral genotype. Accordingly, our eQTL analyses serve mainly to describe CHC– and cirrhosis-dependent regulation (including cell-type effects) rather than to prioritise causal genes. However, we identified eQTLs acting specifically in CHC and HCV-induced cirrhosis and different cell-types, confirming that eQTLs are highly context-specific^72,73^. Most interaction signals were missed by healthy bulk models and appeared only when we modelled cirrhosis and cell-type abundance. Such approaches can help to nominate context-dependent targets for functional follow-up even when GWAS hits do not colocalize with baseline eQTLs^73^. Bulk RNA sequencing does not capture the spatial and cellular complexity of the liver. Therefore, we used a deconvolution approach to estimate cell type composition and identify cell-type specific eQTLs. This analysis revealed a progressive shift in the architecture of the liver during CHC, characterised by increased immune and stromal cell population, with additional epithelial and endothelial remodelling as cirrhosis developed. CHC was marked by an increase of antibody-producing plasma and mature B cells and mononuclear phagocytes/macrophages and altered hepatocyte composition. These changes were more pronounced in, particularly for plasma cells, macrophages, stellate cells, cholangiocytes and periportal endothelial cells. Interestingly, hepatocyte clusters 3 and 4 were lost in all BOSON samples. Cluster 3 hepatocytes are enriched for genes involved in lipid and cholesterol synthesis and are thought to represent the periportal region in mice, while the cluster 4 hepatocytes are characterised by activation of immune, fibrinolytic, and triglyceride biosynthesis pathways, with no clear murine counterpart^52^. On the contrary, hepatocyte-clusters 1 and 6, which may correspond to central veinous and interzonal hepatocytes, were more abundant in BOSON patients. It’s important to note that deconvolution relies on the assumption that reference single-cell signatures accurately reflect cell types in the sample, and it can struggle to estimate rare populations^50^. To mitigate this limitation, we used two single-cell reference signatures, including one that contained both healthy and cirrhotic liver tissue.

Finally, for the first time, our PRS analysis showed that scores derived from non-viral liver disease cohorts are associated with HCV-induced cirrhosis, supporting the existence of shared genetic pathways across different liver disease aetiologies. In addition, the absence of significant interactions with viral factors suggests that these PRS could be broadly applicable to individuals with CHC who are at risk of progressive liver disease. We also found that several PRS were associated with decompensated cirrhosis, indicating their potential utility for stratifying patients according to their likelihood of developing more severe liver disease. Although classical clinical scores^74^ currently outperform genetic ones in predictive accuracy, a key advantage of PRS is that they can be calculated well before symptom onset, enabling early identification of high-risk individuals and timely implementation of preventive strategies. Only one PRS was significantly associated with HCC risk among cirrhosis patients. Interestingly, three of the PRS (PGS000726^23^, PGS004621^75^ and PGS000872^22^) had previously been linked to increased genetic risk for liver cancer. This observation reinforces our genetic findings, suggesting that HCC development in the context of CHC may involve distinct mechanisms compared to both HCV-induced cirrhosis and HCC arising from non-viral causes.

One important limitation of this study is that we focused on disease susceptibility rather than the rate of progression (either from CHC to cirrhosis or from cirrhosis to HCC), for which genetic risk factors can differ^76,77^. The time of infection onset was unavailable for most STOP-HCV patients and difficult to estimate accurately. Most studies assume that infection began at the time of first positive anti-HCV test or patient recall but real infection can predate the estimated date by many years^78^. Similarly, the dates of cirrhosis or HCC diagnosis can be imprecise, especially the latter, which is frequently detected at a late stage^79^. Therefore, time-to-event analyses underestimate duration of infection and bias disease progression rates toward the null^80^. Another limitation is the incomplete data on prior antiviral treatment. Fibrosis regression following a sustained virologic response (SVR) may lead to misclassification of treated patients as non-cirrhotic, potentially introducing reverse causation by falsely attributing protection to certain alleles. In addition, patients who survive long enough to be recruited are more likely to include non-responders carrying unfavourable variants, which can introduce collider bias when survival is conditioned upon recruitment^77^. Finally, sex is a major modifier of liver disease trajectories in chronic viral hepatitis, likely reflecting differences in sex-hormone signalling and immune responses^42^. All our association models were adjusted for sex. However, due to the male predominance in the cohort, female-only strata were underpowered, and the formal tests of sex-specific genetic effects were limited.

In conclusion, this study provides a comprehensive examination of the host genetic factors underlying HCV-induced cirrhosis and HCC and its interaction with HCV genotypes. Our findings highlight a shared genetic architecture for cirrhosis across different aetiologies, while underscoring that the pathways to HCC in the context of CHC are genetically distinct.

## Data availability

All summary statistics are available in the manuscript or at https://doi.org/10.6084/m9.figshare.29877302.

## Acknowledgements

The authors thank HCV Research UK (funded by the Medical Research Foundation) for their assistance in handling and coordinating the release of samples for these analyses. The authors also thank Gilead Sciences for the provision of samples and data from the BOSON clinical study for use in these analyses, and the NHS patients who participated in STOPHCV1 across multiple sites in the UK. This study is funded by a grant from the Medical Research Council (MR/K01532X/1–STOP-HCV Consortium). This study is partly funded by the Cancer Research UK under the DeLIVER (The Early Detection of Hepatocellular Liver Cancer) Project (Award ID C30358/A29725). PK is supported by Wellcome (222426/Z/21/Z) and CRUK (DRCNPG-Nov22/100005). GC is supported in part by NIHR BRC of Imperial College NHS Trust. EB is supported as an NIHR senior Investigator and acknowledges the Oxford NIHR Biomedical Research Centre. MAA is supported by a Sir Henry Dale Fellowship jointly funded by the Royal Society and Wellcome Trust (220171/Z/20/Z).

## Notes

### Competing Interest Statement

The authors have declared no competing interest.

### Author Declarations

The sampling protocols for all cohorts were approved by the appropriate institutional review boards, and all patients provided written informed consent. All studies conform to the ethical guidelines of the 1975 Declaration of Helsinki. The HCVRUK patients were enrolled by consent into the HCV Research UK registry. Ethics approval for HCV Research UK was given by NRES Committee East Midlands – Derby 1 (Research Ethics Committee reference 11/EM/0314). The BOSON study protocol was approved by each institution's review board or ethics committee before study initiation (clinical trial registration number: NCT01962441). For this, we only received and used de-identified individual-level data. All direct identifiers had been removed prior to transfer, study sites were coded, and we had no access to any re-identification key. The STOPHCV1 trial was approved by the Cambridgeshire South Research Ethics Committee (15/EE/0435). The trial was registered at ISRCTN (37915093, 11 th April 2016), and EudraCT (2015-005004-28, 31 st December 2015).

