## Supplementary Figures for "Genome-wide association and gene-virus interaction study of liver disease in hepatitis C virus-infected patients"

**Supplementary Figure 1. Principal components analysis of the STOP-HCV GWAS patients.** Scatter plot of PC1 and PC2. Each point represents one patient, color-coded by self-identified race/ethnicity.

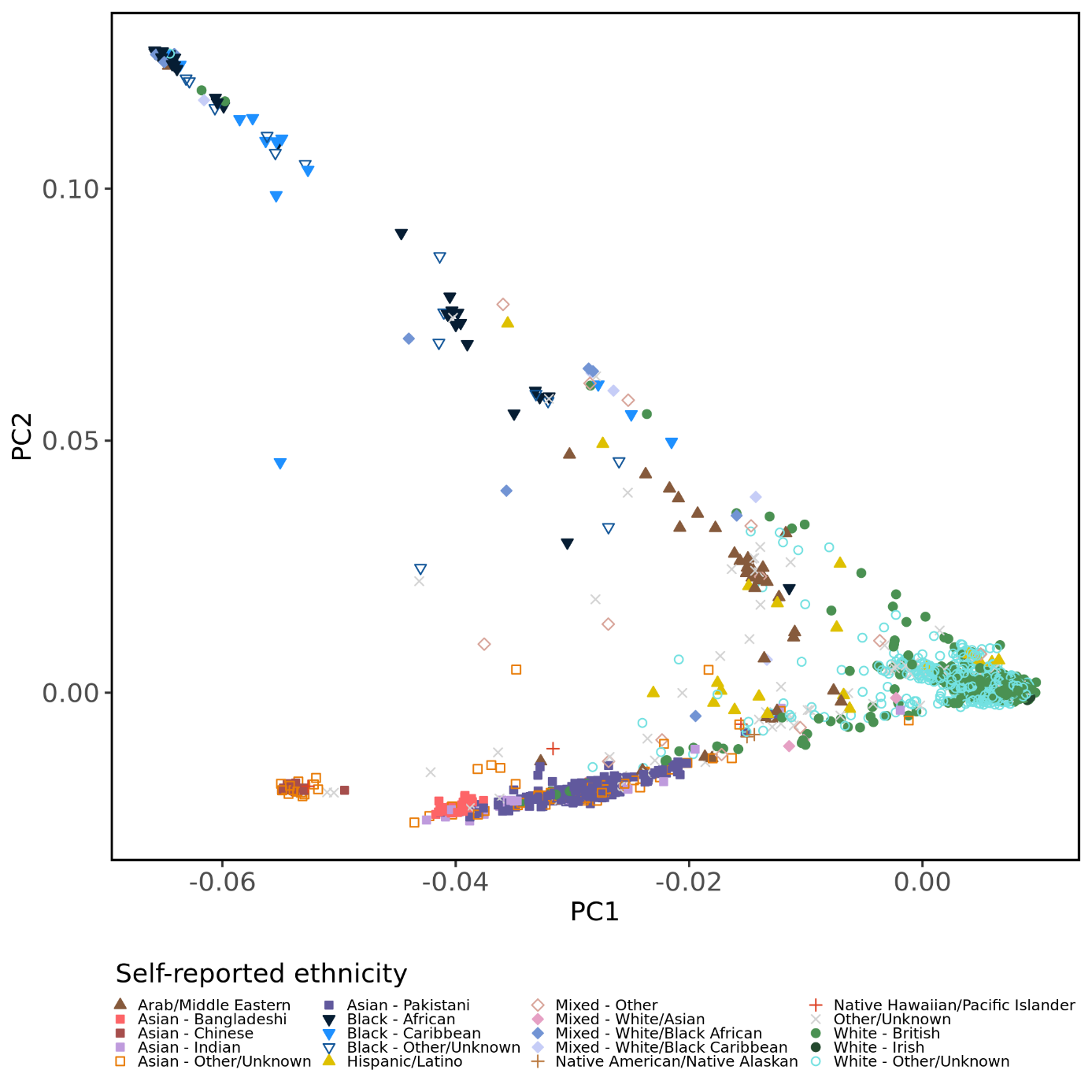

**Supplementary Figure 2. Definition of European samples in the STOP-HCV cohort.** A. Admixture plot of the STOP-HCV cohort and 1000G. Individuals with EUR ancestry > 80% were defined as Europeans. B. Principal components analysis of the European STOP-HCV GWAS patients subgroup after projection onto 1000G phase 3 populations.

| A | 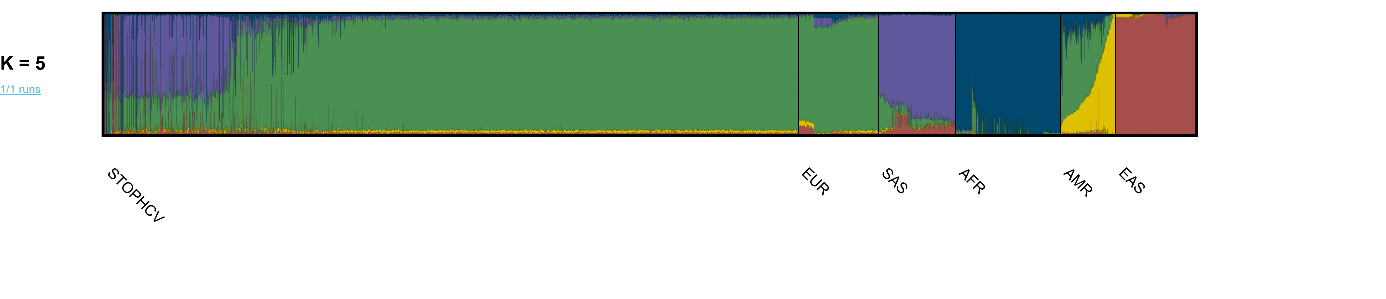 |
| --- | --- |
| B | 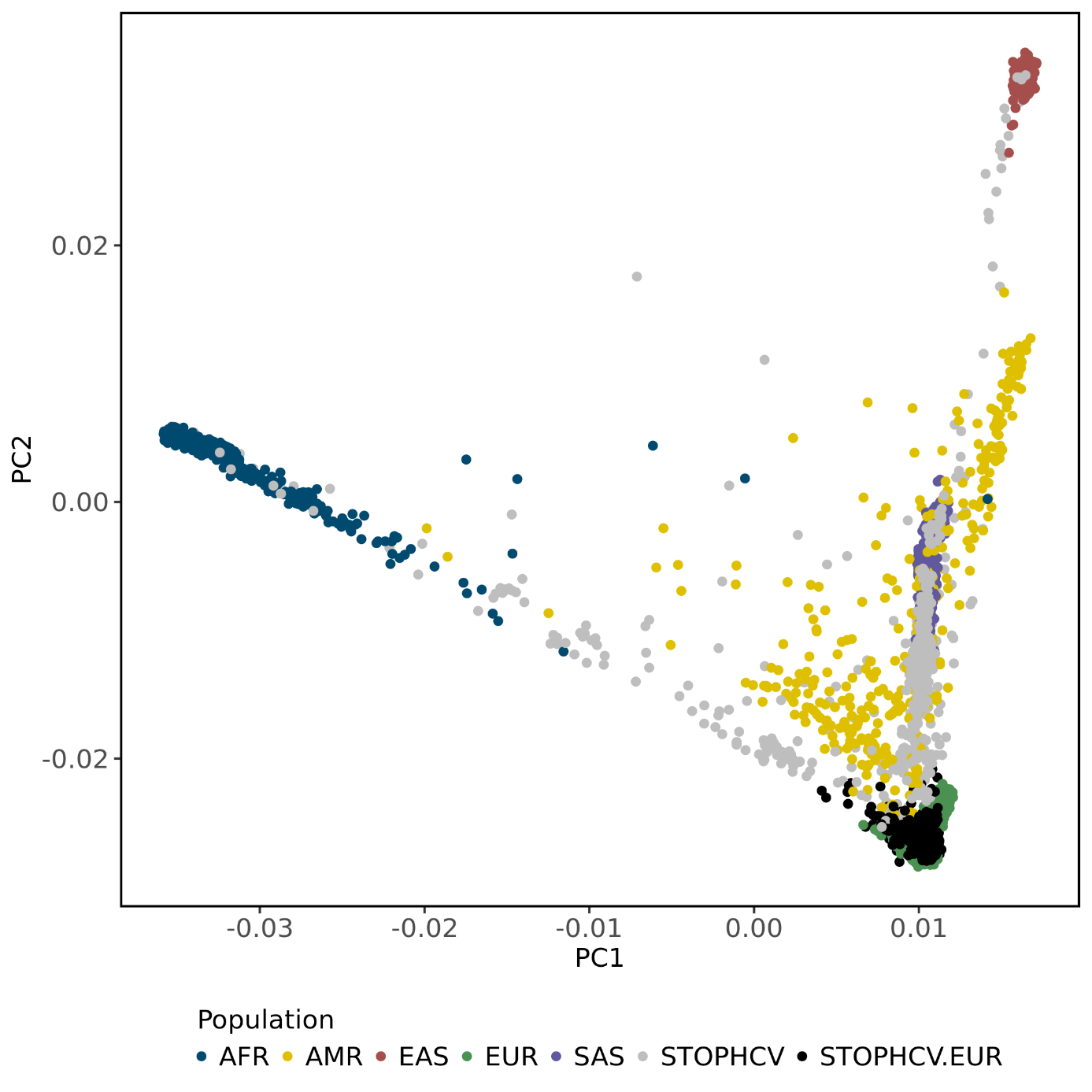 |

**Supplementary Figure 3. Parallel coordinate plot**. PC1 to PC32 were standardized and the value for each STOP-HCV patient was plotted as a line, color-coded by self-identified race/ethnicity. Most of the variation are captured by the first 10 PCs (lines tend to be at the outer ranges of the distribution).

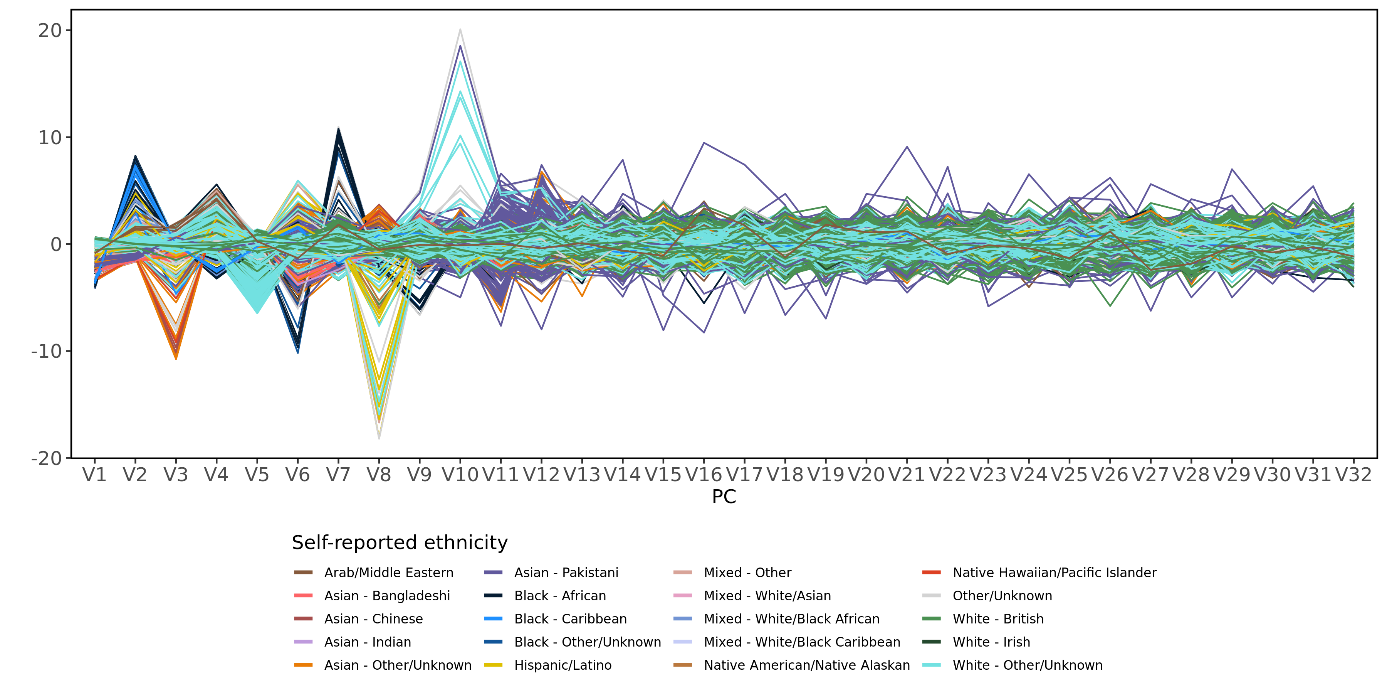

**Supplementary Figure 4**. Principal component analysis on the normalized gene counts matrix.

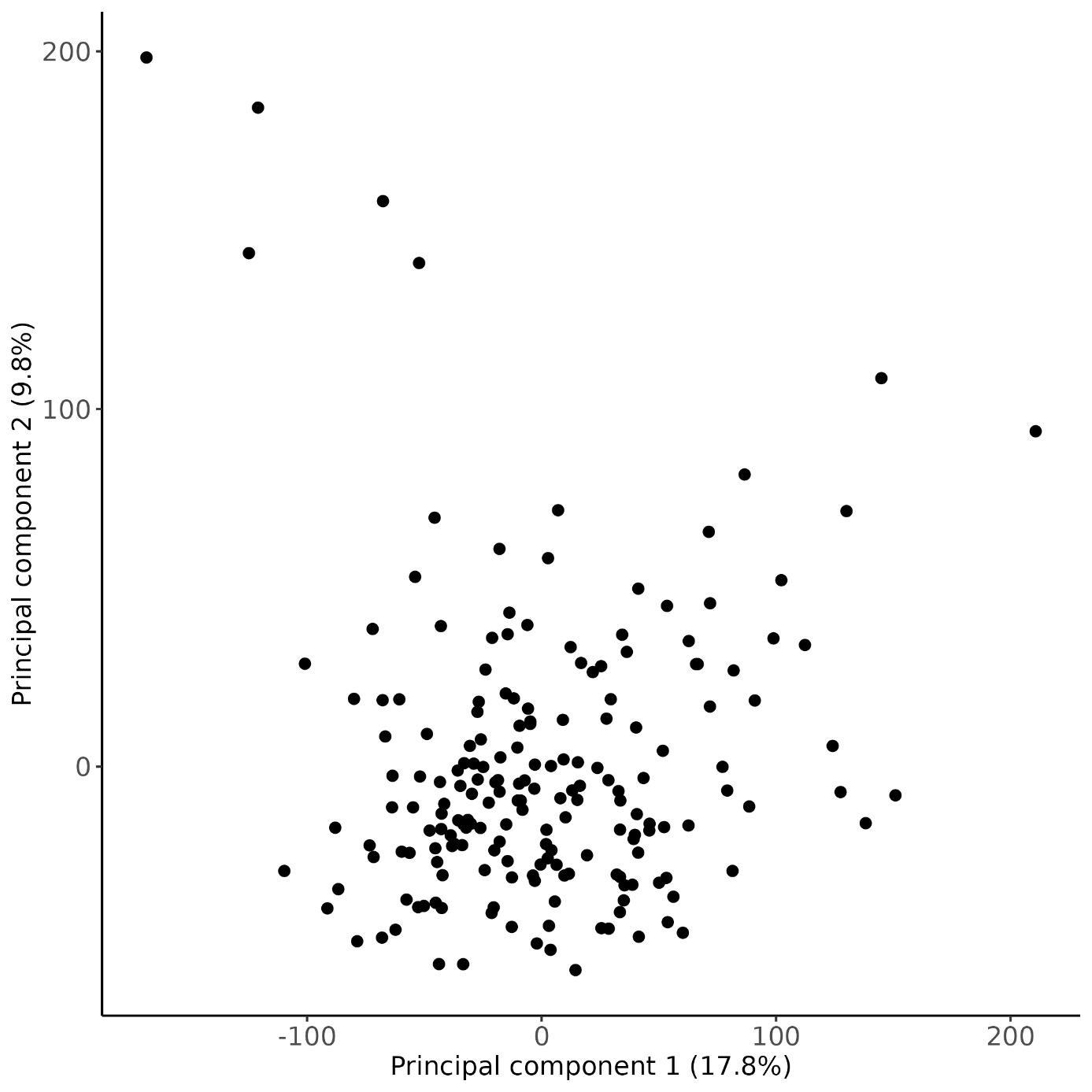

**Supplementary Figure 5. Association between the 10 first gene counts principal components and covariates.** Significant associations are labelled on panel A and the most significant are displayed on panels B to G.

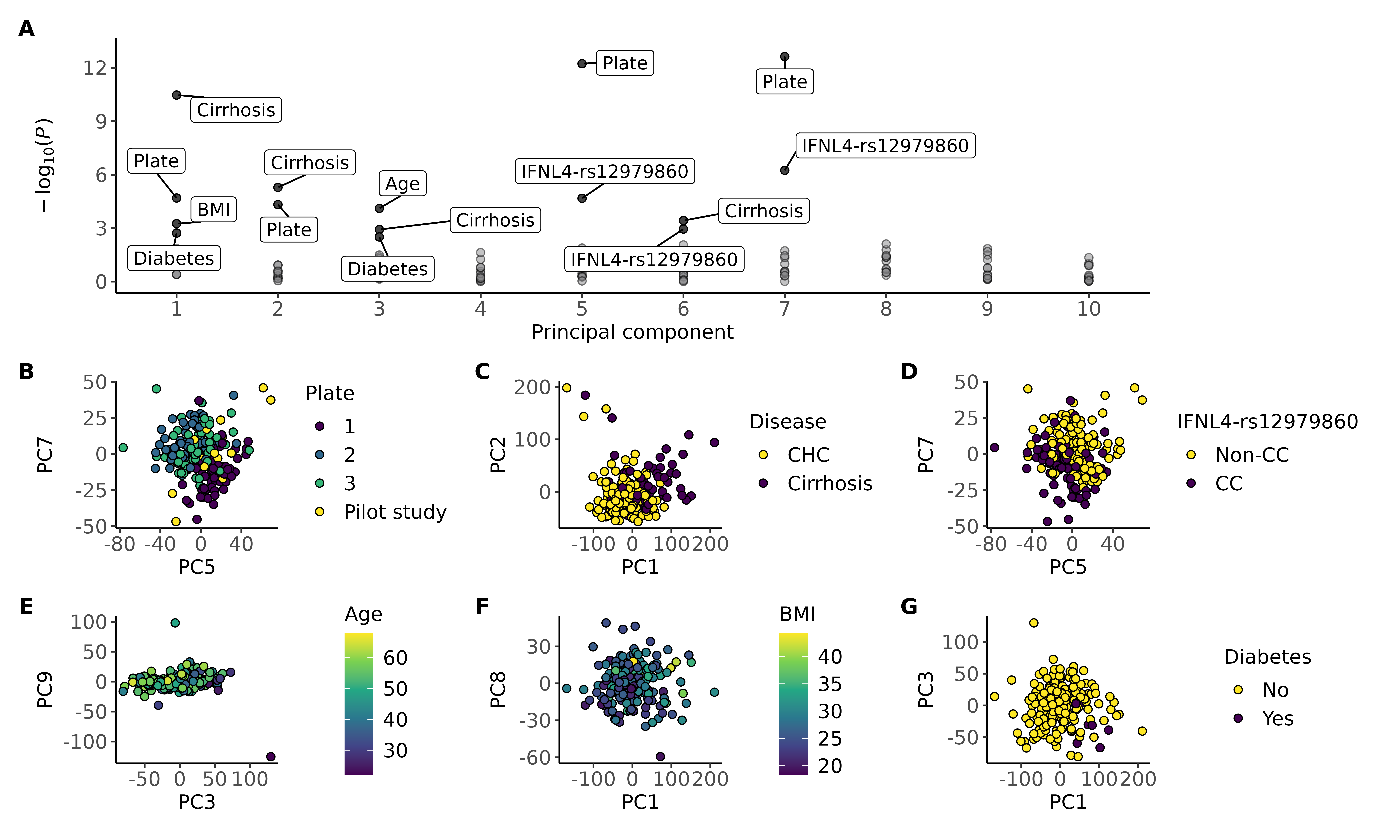

**Supplementary Figure 6**. Quantile-quantile plots for association analyses of genetic

variants with cirrhosis and HCC.

| Cirrhosis vs CHC | HCC vs cirrhosis |  |
| --- | --- | --- |
| 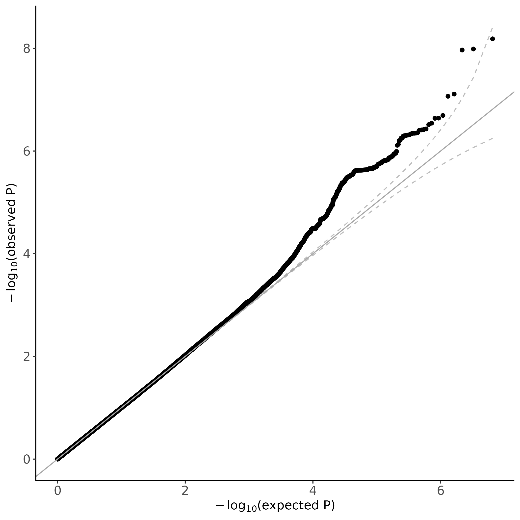 | 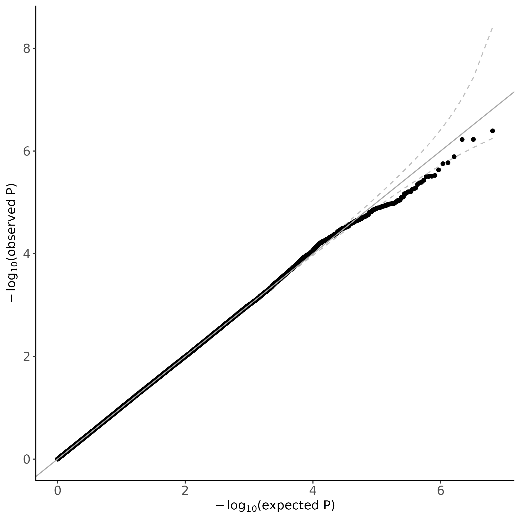 | All patients |
| 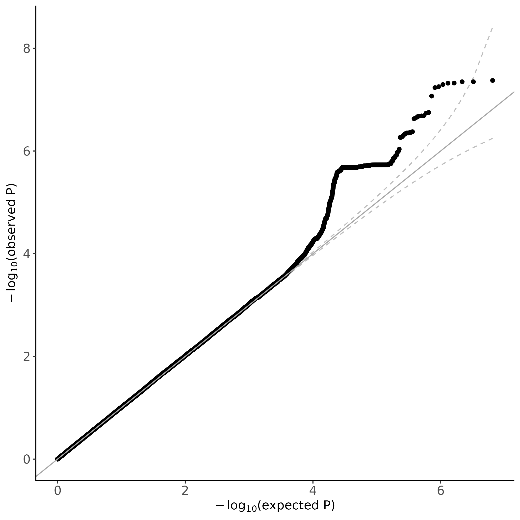 | 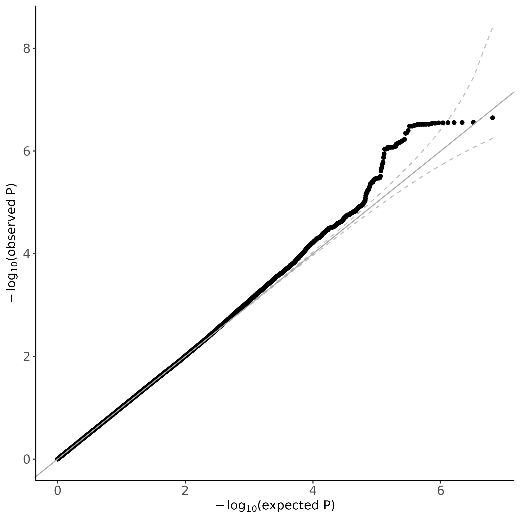 | Gt1 |
| 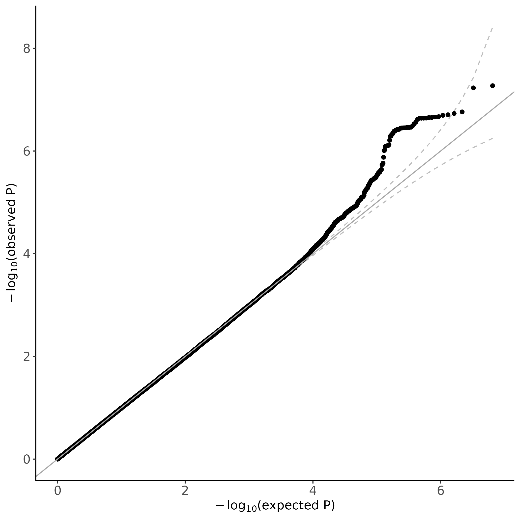 | 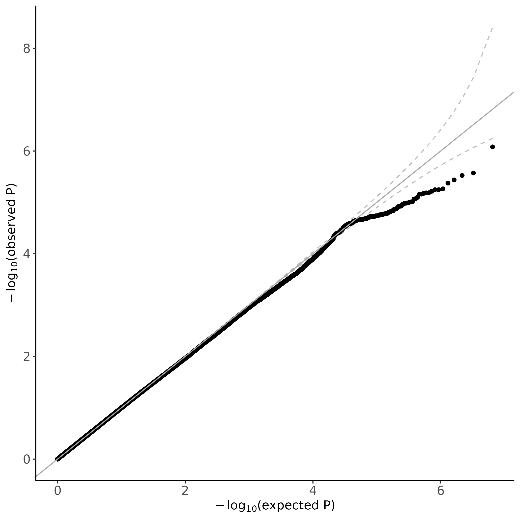 | Gt3 |

**Supplementary Figure 7. GWAS of HCC using all non-HCC patients (with and without cirrhosis) as controls.** A. Manhattan plot. B. QQ plot.

| A | 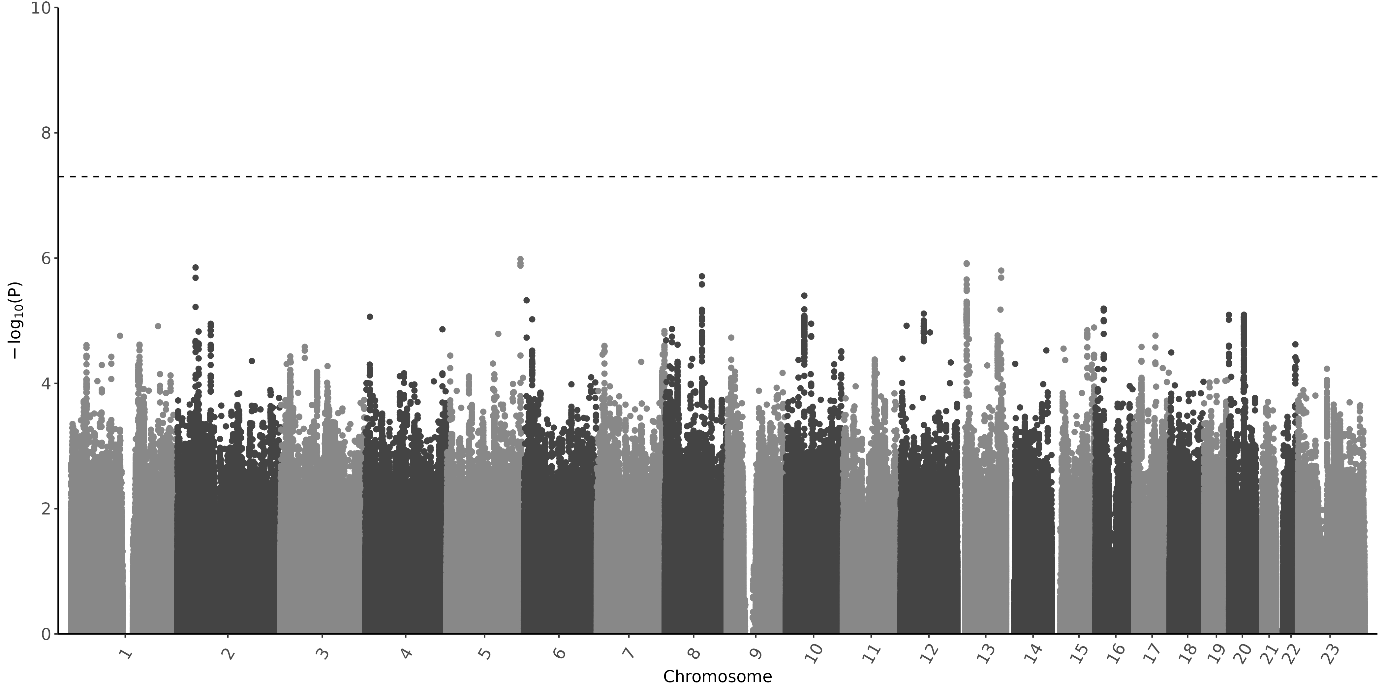 |
| --- | --- |
| B | 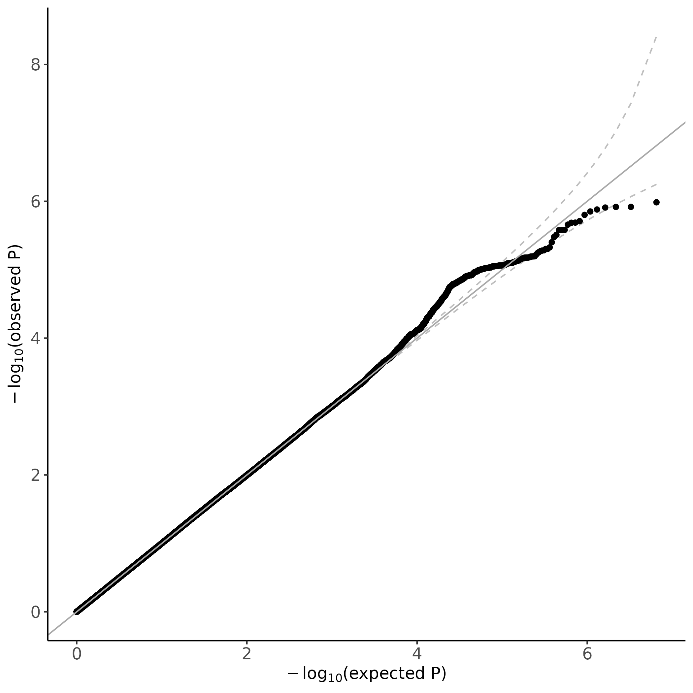 |

**Supplementary Figure 8**. Forest plot for the previously reported HCC risk variants tested in the STOP‑HCV cohort.

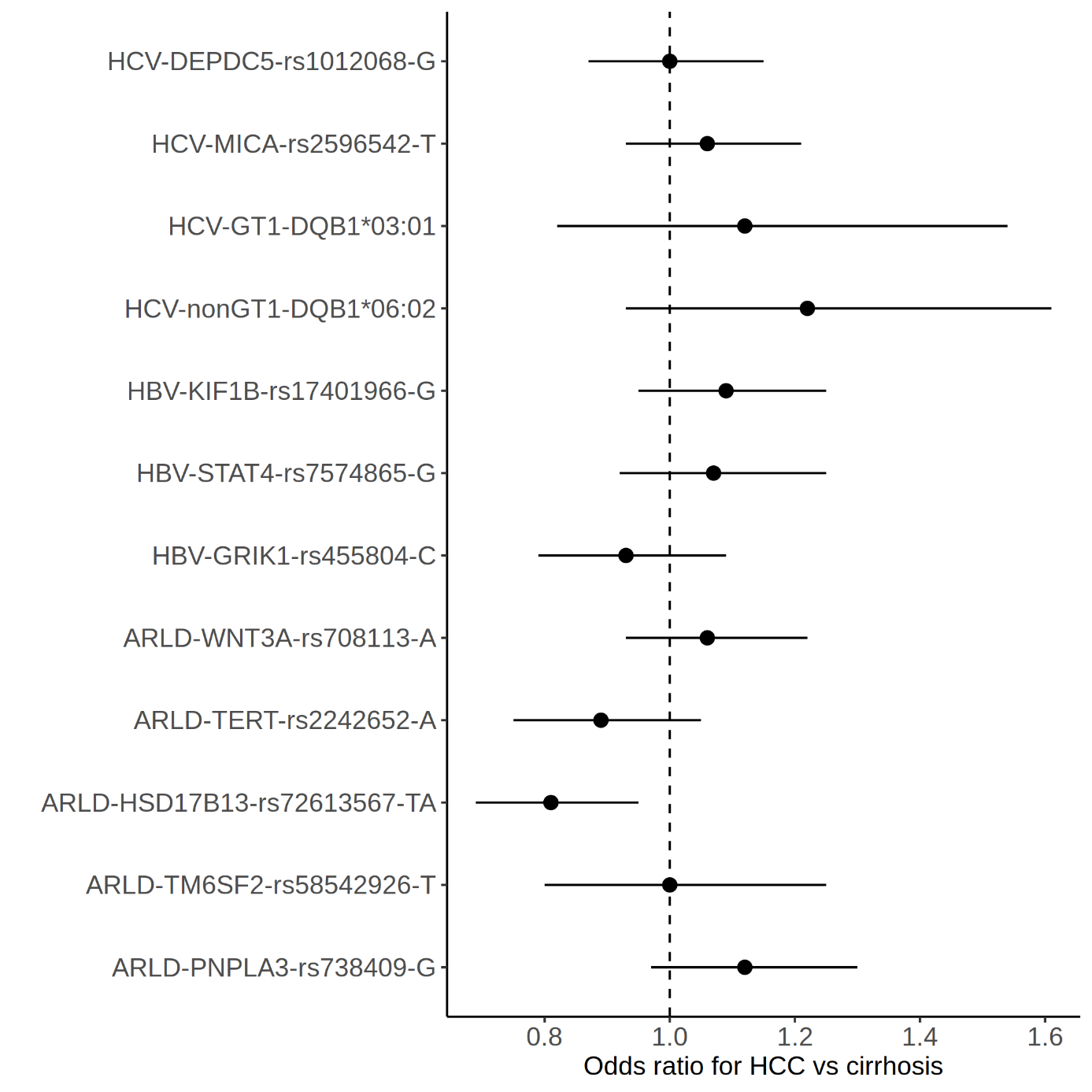

**Supplementary Figure 9.** Regional plots for the NT5DC1/COL10A1 and SUGP1 loci close to genome-wide significance in gt3 patients.

A

B
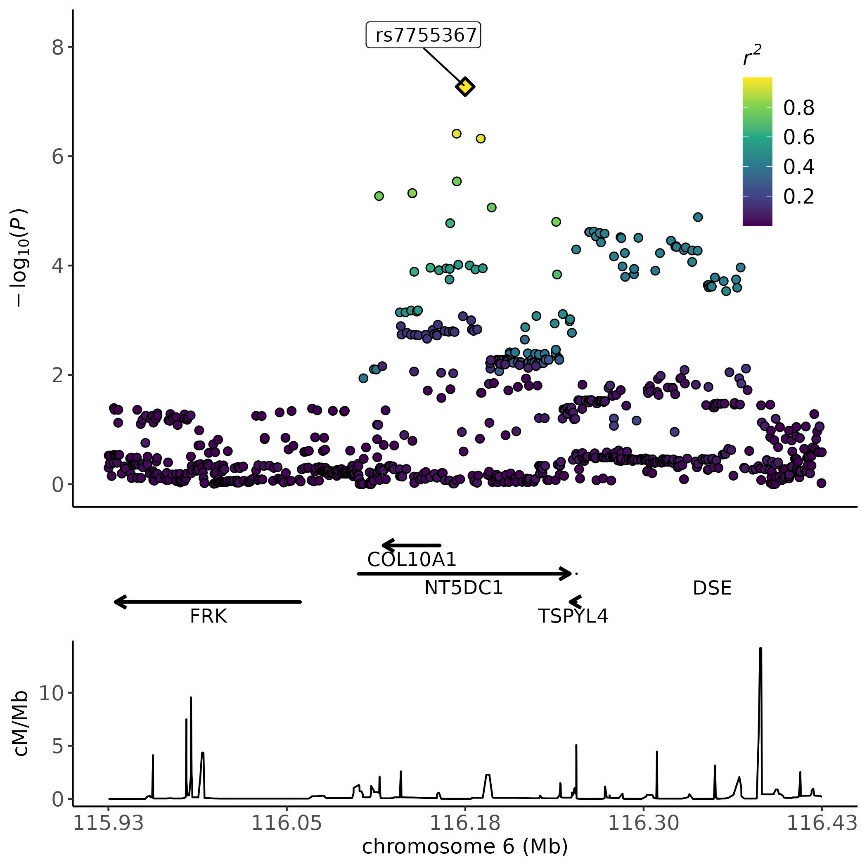

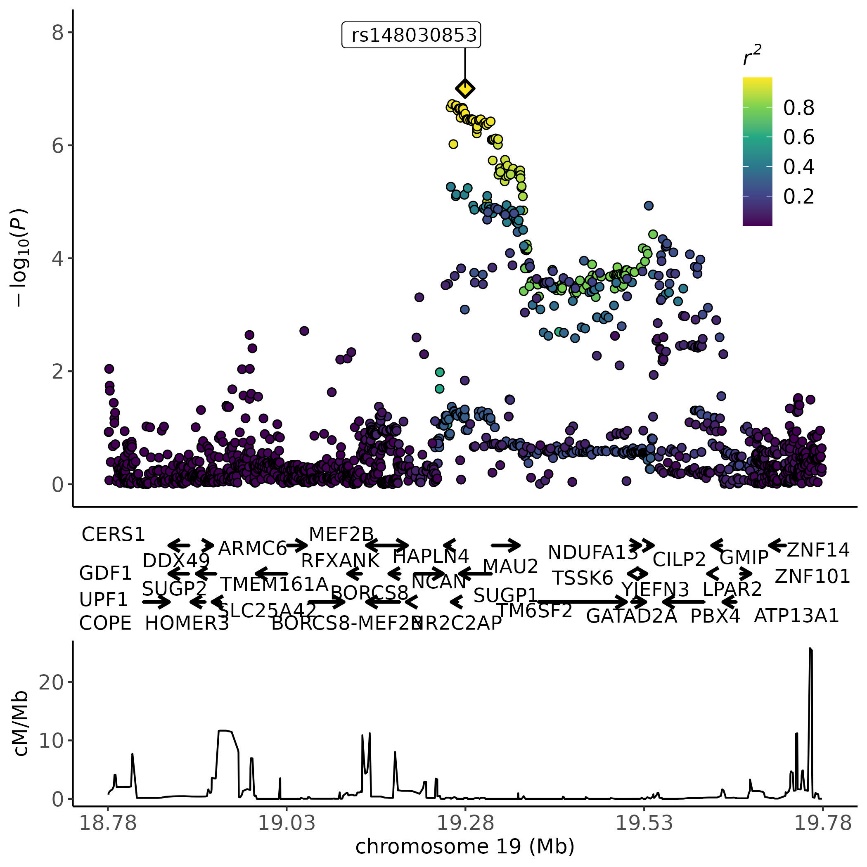

**Supplementary Figure 10. Top view of the 3-dimensional structure of HLA-DR (left) and HLA-DQ proteins (right),** based on PDB (accession codes: 1A6A and 1JK8) with the DR/DQ α chains shown in yellow and the DR/DQ β chain in purple. Amino acid position identified by the association analysis within the HLA-DRβ1 protein is shown for position 37 (cyan) and within HLA-DQβ1for position 45. Peptide binding is shown in green.

| HLA-DRβ1 position 37 | HLA-DQβ1 position 45 |
| --- | --- |
| 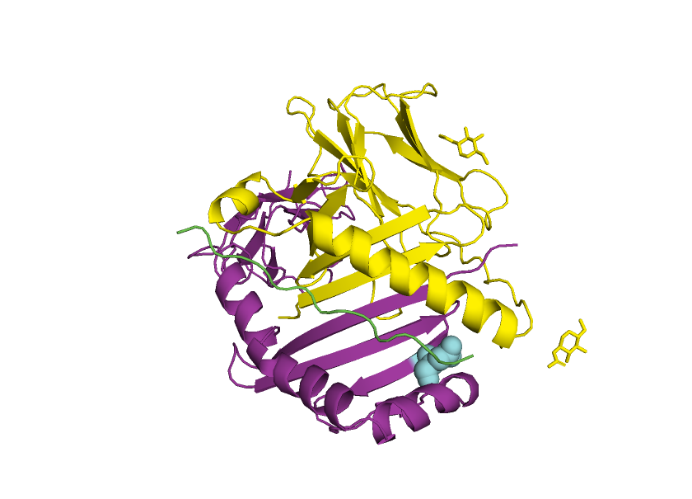 | 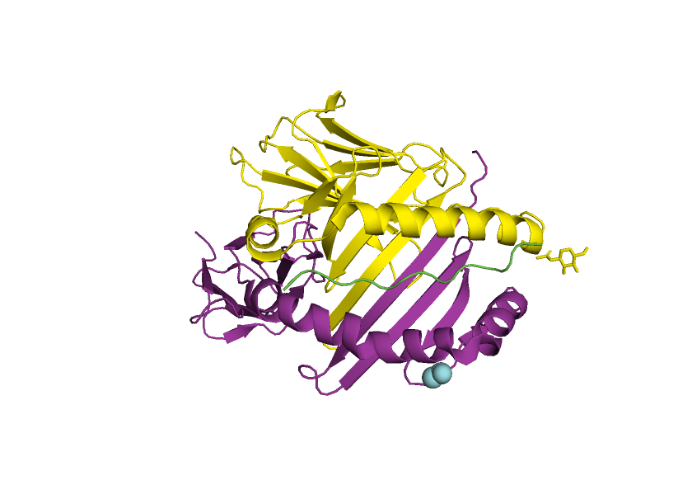 |

**Supplementary Figure 11.** Proportion of eGenes from each GTEx tissue that overlapped with BOSON eGenes.

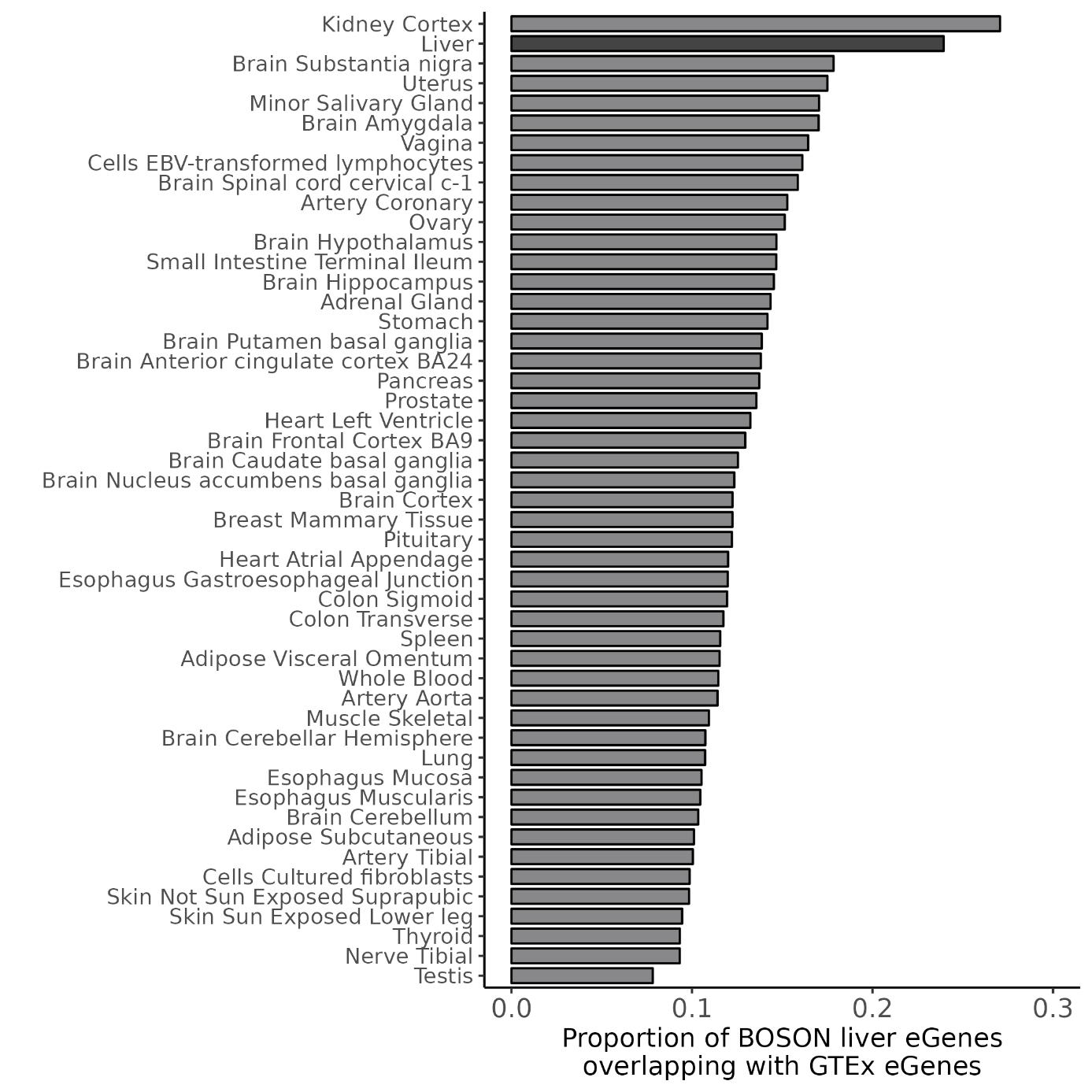

**Supplementary Figure 12.** Gene expression levels of BOSON eGenes, cirrhosis ieGenes and non-eGenes in BOSON top panel) and GTEx datasets (bottom panel).

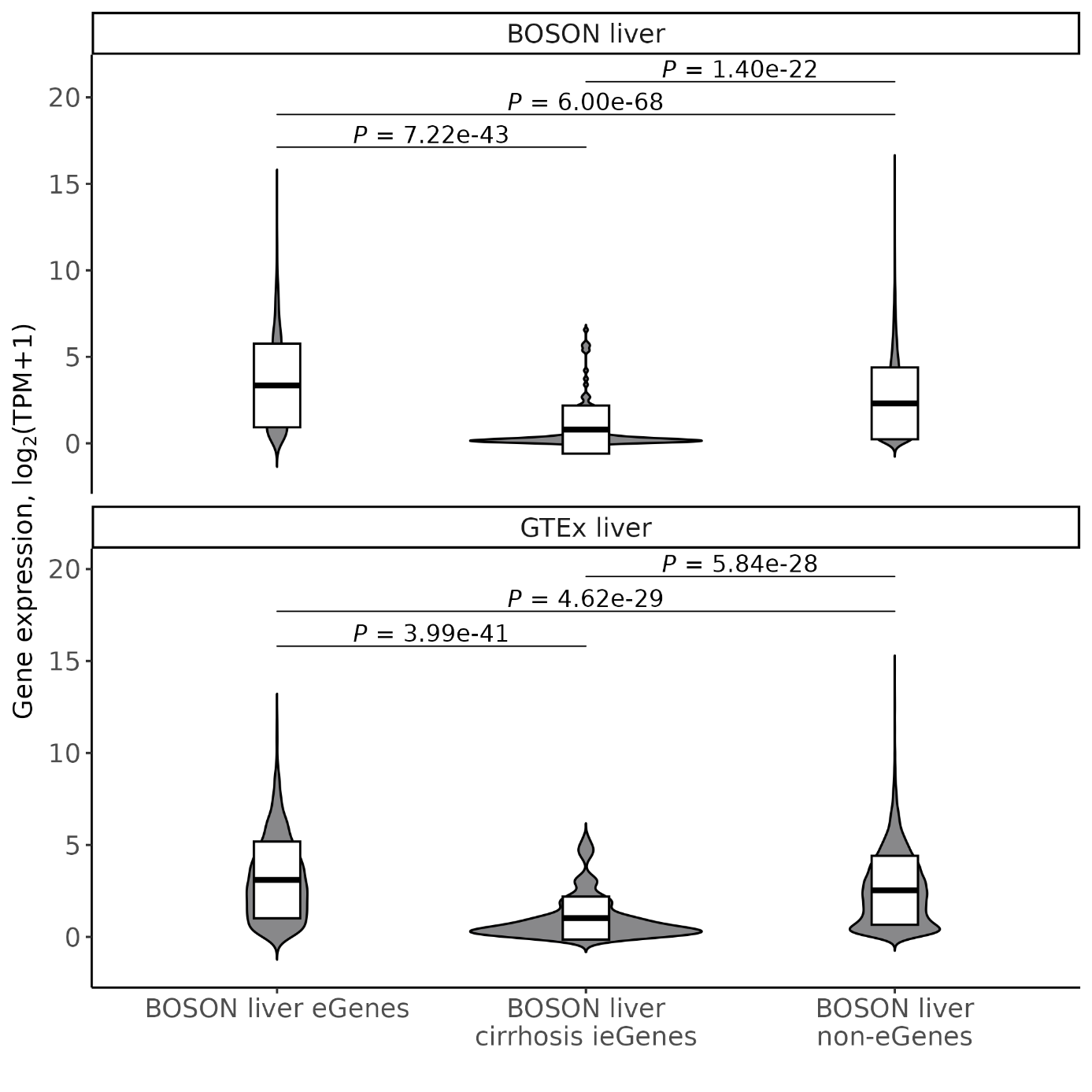

**Supplementary Figure 13.** Scatter plot of the effect sizes (β/se or Z-scores) of BOSON eQTLs (left panel) and cirrhosis ieQTLs (right panel) with GTEx eQTLs for BOSON eGenes shared with GTEx (top panel) and non-shared with GTEx (bottom panel).

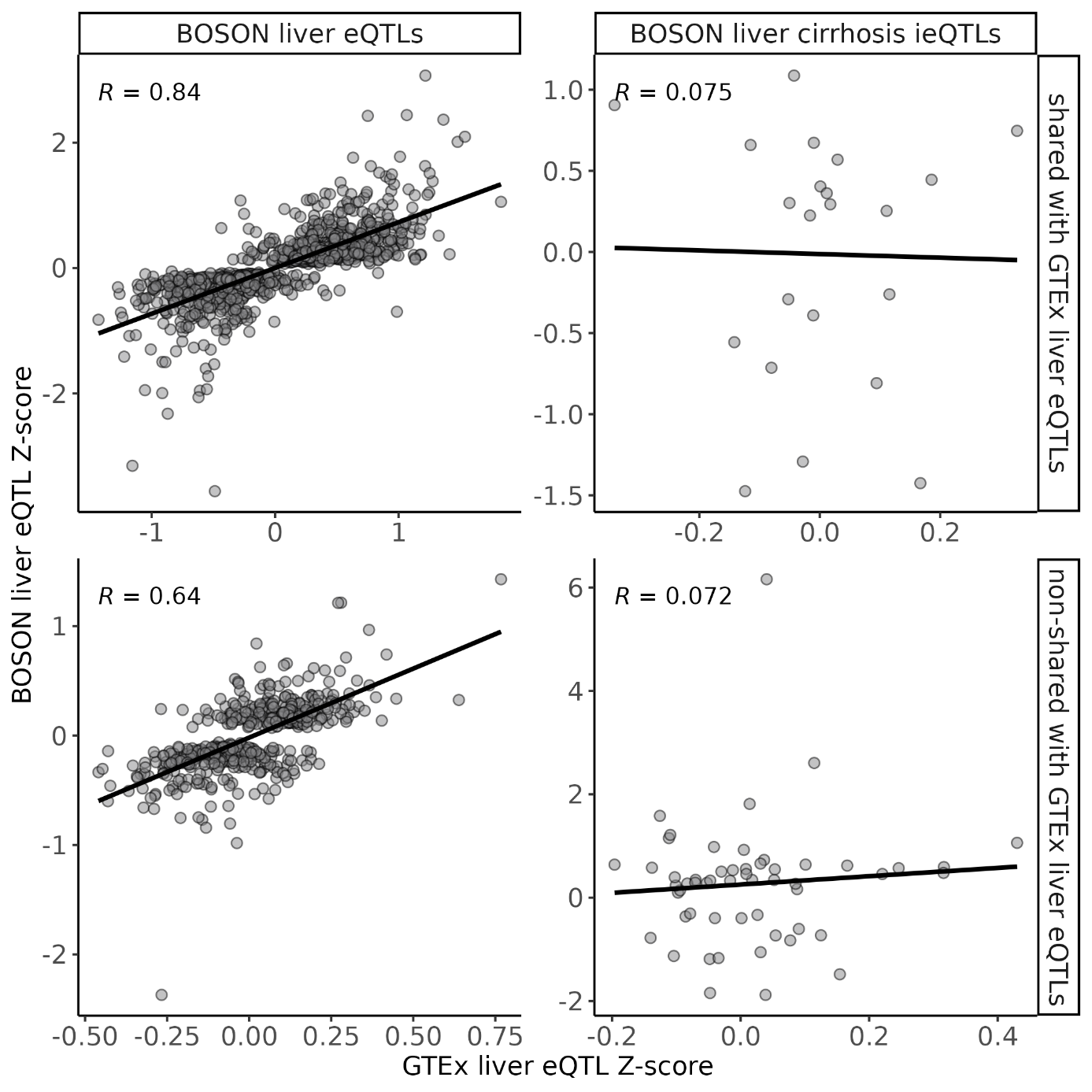

**Supplementary Figure 14.** Estimated cell type proportions in BOSON and GTEx bulk liver RNA-seq using signature matrices from MacParland et al (healthy liver) and Ramachandran et al (healthy and cirrhotic liver).

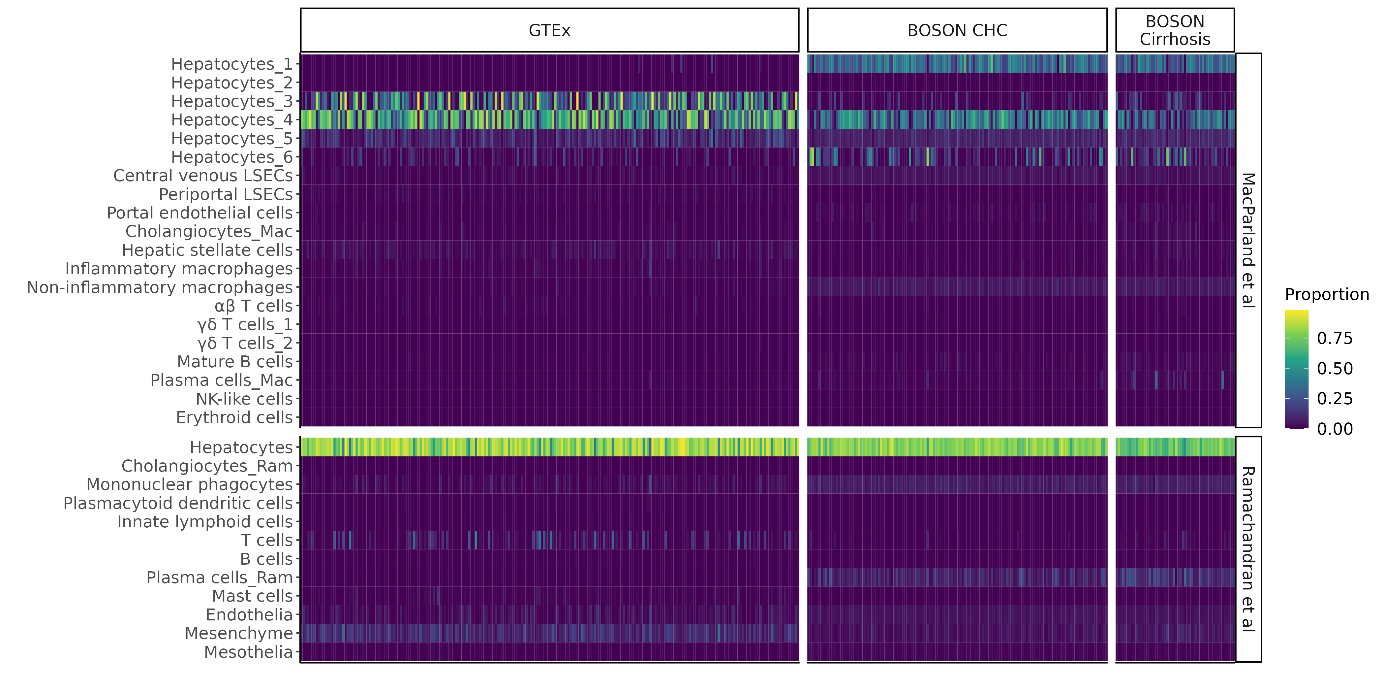

**Supplementary Figure 15.** Scatter plot of BOSON cell-type significant ieQTLs effect sizes (β/se or Z-scores) and the corrresponding bulk eQTL effect size, among the cell-type ieQTLs shared (right panel) or non-shared (left panel) with the bulk eQTLs.

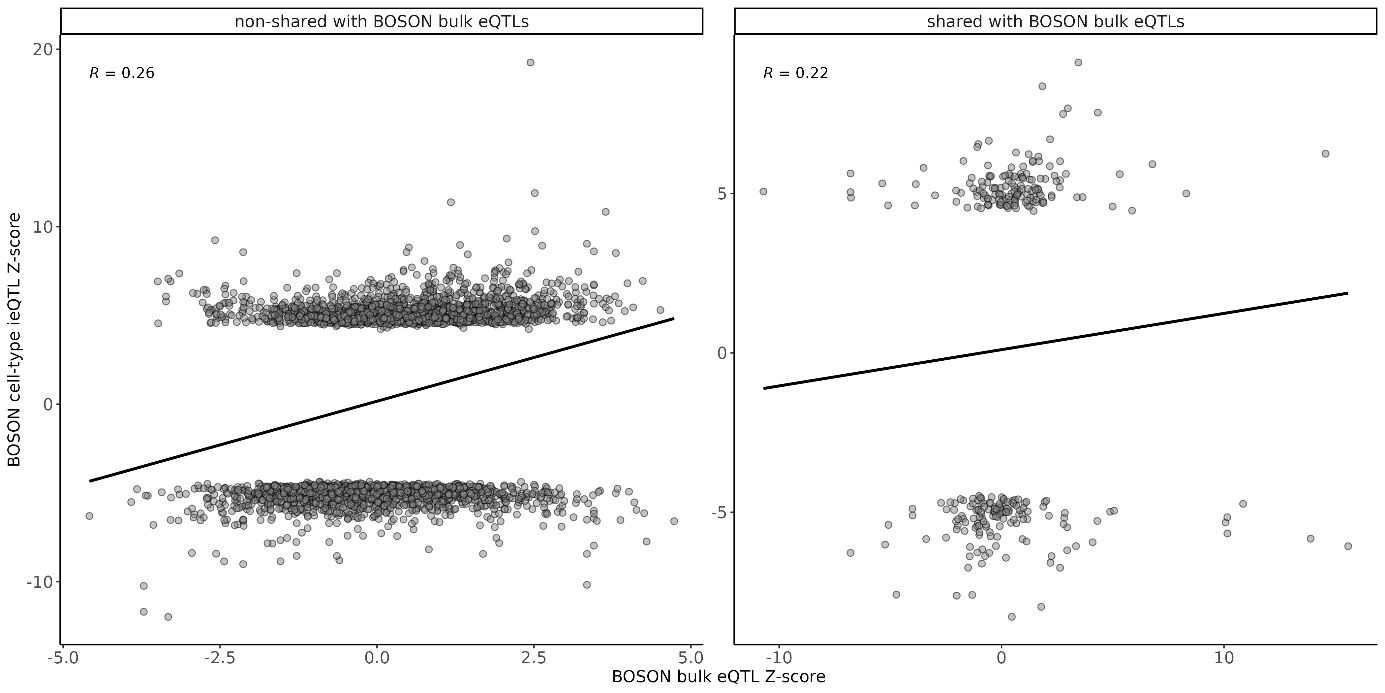
